# Epidemiological, vectorial and landscape changes in the context of declining *Onchocerca volvulus* transmission across the Kakoi-Koda focus, Ituri, Democratic Republic of the Congo

**DOI:** 10.64898/2026.03.19.26348782

**Authors:** Luís-Jorge Amaral, Tony Ukety, Jules Upenjirwoth, Deogratias U. Wonya’Rossi, Michel Mandro, Francoise Nyisi, Pascal Adroba, Wilma A. Stolk, Joseph N. Siewe Fodjo, María-Gloria Basáñez, Anne Laudisoit, Robert Colebunders

## Abstract

**Background:** Onchocerciasis remains a public-health challenge in the Democratic Republic of the Congo (DRC). The Kakoi-Koda focus, Ituri Province, exhibited high endemicity in the early 2000s and received community-directed treatment with ivermectin (CDTI) in some health zones (e.g., Nyarambe), but not in others (e.g., Logo). Moxidectin clinical trials were conducted in these health zones, alongside onchocerciasis-associated epilepsy studies.

**Methodology:** We synthesised epidemiological (including nodule prevalence), entomological and CDTI programmatic data. We collated anti-Ov16 serological data from epilepsy-related studies (community, cohort, case-control designs, 2015-2021) and skin-snip microscopy results from two moxidectin trial screenings (2009-2011; 2021-2023) and epilepsy-related studies (2015-2017). Geospatial analyses were used to describe land-cover change relevant to vector ecology and to identify areas with recent transmission.

**Principal findings:** Onchocerca *volvulus* transmission declined markedly over time. In CDTI-naïve Logo villages, microfilarial prevalence fell from 69-79% (first trial, 2009-2011) to 9% (second trial, 2021-2023), and mean infection intensity from 17-26 to 1 microfilariae per skin snip, similar to declines observed in Nyarambe villages under CDTI (72% to 3% and 11 to 0.4, respectively). Anti-Ov16 seroprevalence among children aged 3-10 years was low (0-5%) from 2016 onwards, and seropositivity was geographically circumscribed, mirroring contemporary skin-snip results. Human landing catches and breeding-site prospections (2015-2017) identified *Simulium dentulosum* and *S. vorax* as the current anthropophagic species, with no evidence of *S. neavei* after 2009. Progressive deforestation and canopy opening provide a plausible mechanism for a shift from crab-associated *S. neavei* habitats towards more open-habitat vectors.

**Significance:** Consistent parasitological, serological, entomological and geospatial evidence indicates substantially reduced transmission across Kakoi-Koda, with spatially-circumscribed residual transmission. Whether the current simuliid species can sustain transmission above elimination thresholds remains uncertain. Targeted, integrated surveillance is warranted to guide CDTI and stop-CDTI decisions. The dataset assembled here can be used to inform transmission modelling of these dynamics.

**Author Summary:** Onchocerciasis, also known as river blindness, is a parasitic disease of public health concern in sub-Saharan Africa, transmitted by blackfly vectors. The disease is responsible for skin and eye clinical manifestations and is associated with neurological complications. We investigated an area in north-eastern Democratic Republic of the Congo called the Kakoi-Koda onchocerciasis focus, where the infection was once common. We reviewed and assembled data from past studies on infection in humans and blackflies, and analysed satellite imagery to assess the loss of tree cover that can affect where blackflies live and breed. We found that the prevalence of onchocerciasis in Kakoi-Koda has declined markedly in recent years. This decline appears linked to the disappearance (by deforestation) of the habitat suitable for some blackfly species, and to ivermectin distribution to treat onchocerciasis in parts of the focus. Our findings help to understand why onchocerciasis has decreased across the Kakoi-Koda focus and highlight a small number of fast-flowing river segments where other blackfly species may allow small pockets of local transmission. These results support continued, targeted monitoring to determine whether the disease is still transmitted in specific locations where elimination interventions may be needed.

## 1. Introduction

Onchocerciasis is a neglected tropical disease (NTD) targeted for interruption (elimination) of transmission by the World Health Organization [1]. The causative agent, the filarial nematode worm *Onchocerca volvulus*, is transmitted among humans by female *Simulium* blackfly bites [1]. About 99% of infections occur in sub-Saharan Africa [2], where control relies mainly on community-directed treatment with ivermectin (CDTI) delivered once or twice a year to populations at risk [3]. Ivermectin is primarily microfilaricidal (kills microfilariae, the embryonic stage of the parasite) and transiently suppresses embryogenesis in adult female worms [3]. In addition, a moderate permanent sterilising effect has also been described [4]. Although macrofilaricidal activity (killing of adult worms) has been reported and quantified [5, 6], adult worms can persist for many years despite treatment [3, 7]. Therefore, CDTI must be regularly administered for prolonged periods, accounting for both the long reproductive lifespan of adult worms [8] and the possibility of continued reinfection in areas of intense transmission. Historically, vector control (against blackflies) has also been implemented [3]. The disease is characterised by a range of cutaneous and ocular clinical manifestations (including irreversible blindness), and has been epidemiologically associated with neurological sequelae, such as nodding syndrome, within the spectrum of onchocerciasis-associated-epilepsy (OAE) [9, 10].

Onchocerciasis was first detected in 1902 in the Congo Basin, along the Uélé River in the Democratic Republic of the Congo (DRC) [11]. Since then, several *O. volvulus* foci have been described in the country along major fast-flowing rivers that harbour blackfly breeding sites [11-14]. The African Programme for Onchocerciasis Control (APOC, 1995–2015) supported the scale-up of CDTI in those areas where onchocerciasis was considered to be a public health problem (i.e., where the baseline infection level was at least mesoendemic) [15]. In the DRC, CDTI implementation began in 2001 in Kasai Province, gradually expanding to cover nearly 90% of meso- to hyperendemic villages (i.e., those with a pre-control microfilarial prevalence ≥40% in all ages or a palpable nodule prevalence ≥20% in adult (men) samples) through 22 CDTI projects by 2012 [16-19]. Despite this geographical expansion, therapeutic coverage has been spatially and temporally inconsistent, leaving several areas untreated or only recently treated [20].

The Kakoi-Koda onchocerciasis focus lies on the steep highland slopes of Ituri Province above Lake Albert [17]. In 2003, rapid epidemiological mapping of onchocerciasis (REMO) surveys documented onchocercal nodule prevalence above 40% in several villages of the Kakoi-Koda focus situated in Logo and Nyarambe Health Zones (HZs) [21]. As the remainder of both HZs was considered hypoendemic for onchocerciasis (i.e., <20% nodule prevalence) [18, 19, 22], neither was included in the CDTI Project Ituri North launched in 2007–2008 [22]. Between 2009 and 2011, villages in Logo and Nyarambe around the Kuda and Lebu River basins, where CDTI had never been implemented, were selected for a Phase III randomised, double-blind, single-dose clinical trial comparing the efficacy of moxidectin to that of ivermectin in reducing and suppressing *O. volvulus* microfilarial load (3110A1-3000) [23, 24]. In 2009–2011, the trial enrolled participants aged ≥12 years with at least ten *O. volvulus* microfilariae per milligram of skin and without *Loa loa* or lymphatic filariasis microfilaraemia, confirming high prevalence and substantial microfilarial loads in the focus [23]. After the trial, CDTI was not initiated in Logo, whereas Nyarambe received CDTI for lymphatic filariasis control [20].

In 2021–2023, a second moxidectin clinical trial (MDGH-MOX-3001 and 3002) screened individuals living in the same HZs [21]. Few individuals met the 2009–2011 original inclusion criterion of at least ten microfilariae per milligram of skin, prompting the screening of additional villages within the two HZs in the Awo River basin and the relaxation of the enrolment criterion to include participants with at least one microfilaria in their two skin snips [21]. Comparison of the skin snip screening data between the original Phase III trial (2009–2011) and the subsequent Phase IIIb (2021–2023) trial indicated a marked reduction in *O. volvulus* infection prevalence and intensity across the study area, including in CDTI-naïve Logo HZ villages with comparable sample sizes in both periods [21].

Given the high baseline onchocerciasis endemicity in the Kakoi-Koda focus and its subsequent decline despite inconsistent control efforts, it became important to clarify the fine-scale eco-epidemiological drivers of this change in transmission conditions, particularly for the purposes of analysing the results of the Phase IIIb trial in terms of moxidectin efficacy and effectiveness compared to ivermectin. In this study, we synthesise available epidemiological, (anti-Ov16) serological and entomological data from the Kakoi-Koda focus, and relate these to ecological factors such as forest-cover change around streams in the last 25 years.

## 2. Methods

### 2.1 Ethical statement

This study was conducted under approvals from the Ethics Committee of the University of Antwerp (B300201525249; B300201733350), the Ethics Committee of Ngaliema Hospital, Kinshasa (P: Eth/436/2015) and the Ethics Committee of the University of Kinshasa School of Public Health (ESP/CE/013/2018). Permissions were also obtained from provincial and health-zone authorities.

### 2.2 Study setting and periods

The demographics of the Kakoi-Koda focus are presented according to the current territorial and administrative division of health, namely territories, health zones (HZs) and health areas (HAs). The study area is located in the Ituri Province of the DRC (Mahagi and Djugu territories), in the five HZs comprising the Kakoi-Koda onchocerciasis focus, namely Angumu, Logo, Mahagi, Nyarambe and Rethy (Fig 1). The focus occupies approximately 700 square kilometres along the northeastern Ituri Highlands escarpment from the Blue Mountains near Mount Aboro down towards Lake Albert [17]. Elevation ranges from roughly 620 to 2,450 metres above sea level. The focus lies largely between the Kakoi and Koda rivers, with the Kuda, Lebu and Awo rivers crossing it (Fig 2) [17].

**Fig 1.**
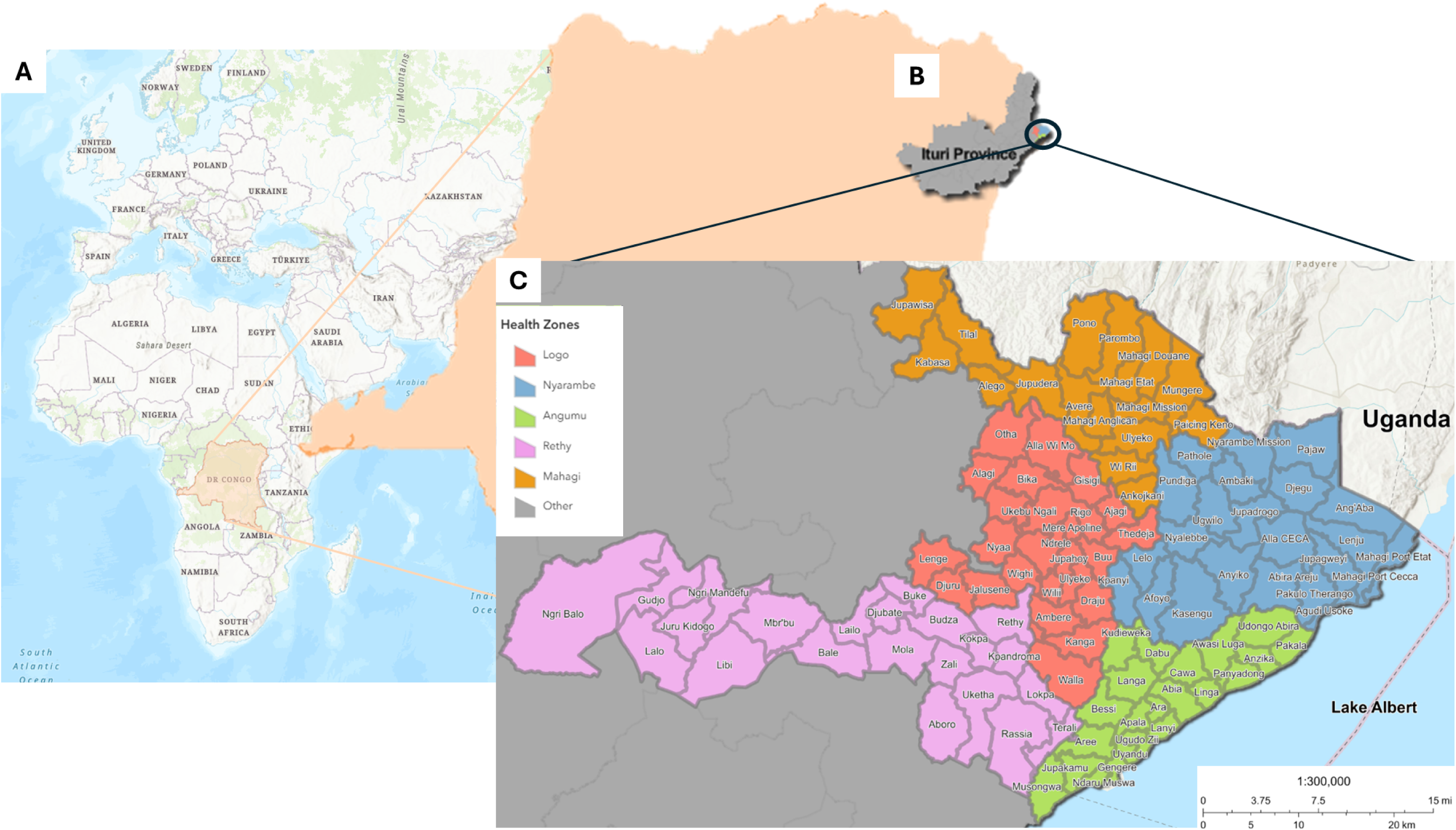
Location of the study areas in the Democratic Republic of the Congo (DRC). (A) Position of the DRC (orange) within Africa. (B) Ituri Province (dark grey) highlighted within the DRC. (C) Map showing the Health Zones (HA) of Angumu (green), Logo (blue) and Nyarambe (red) in East Ituri Province, with their respective Health Areas (HA) delineated (light grey). The map was produced in ArcGIS Pro (Esri, Redlands, CA, USA) using publicly available spatial datasets (GRID3 COD – Health Areas v5.0 [27] and GRID3 COD – Health Zones v5.0 [28]).

**Fig 2.**
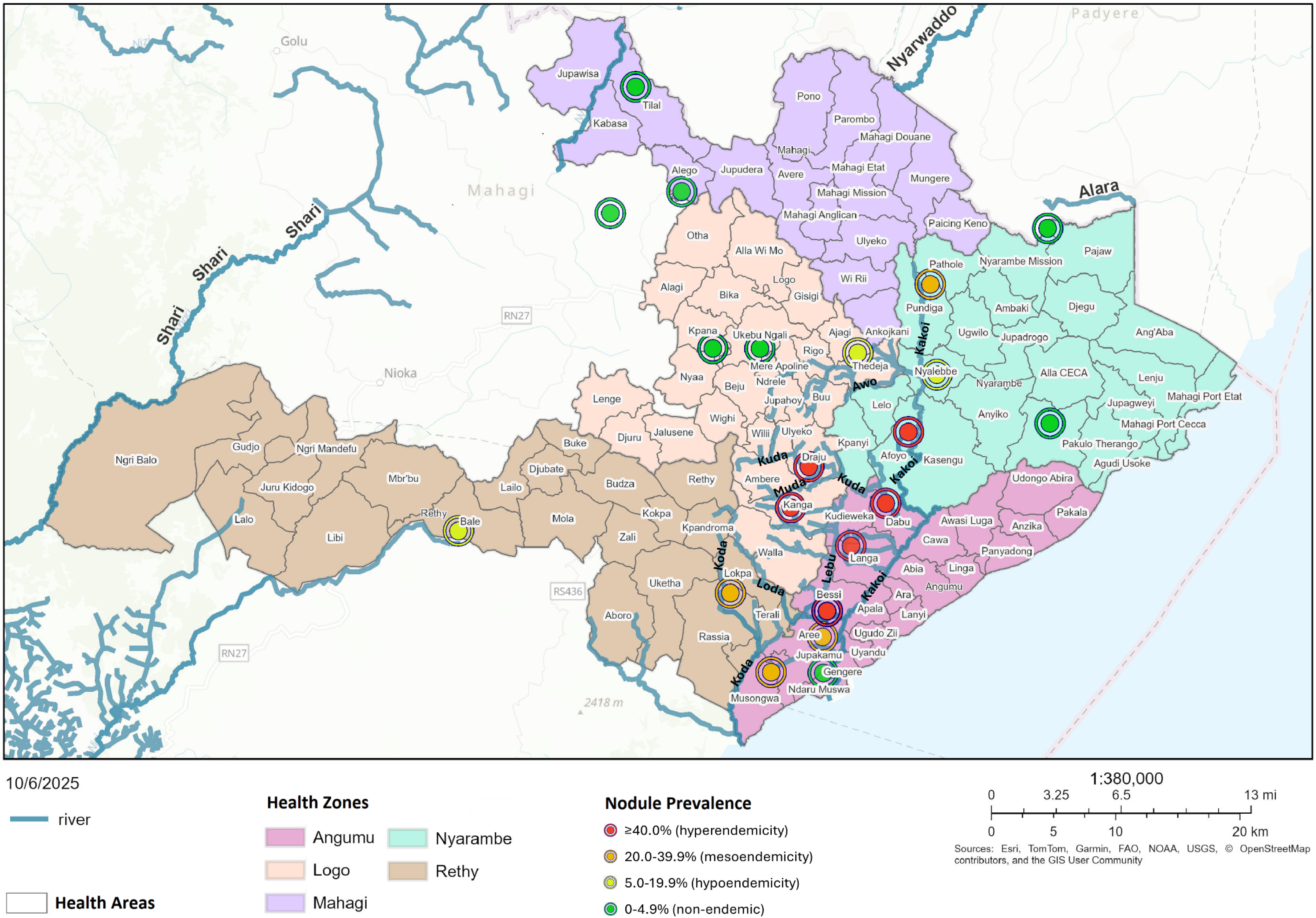
Nodule prevalence in the Kakoi–Koda onchocerciasis focus assessed in 2003 by Rapid Epidemiological Mapping of Onchocerciasis (REMO)/Rapid Epidemiological Assessment (REA) [48]. Clusters of ≥20% nodule prevalence delineate the focus in orange (mesoendemicity) and red (hyperendemicity). Areas with <20% nodule prevalence are shown as yellow (hypoendemicity) and green (non-endemic/sporadic endemicity). Only river basins pertinent to delineating the focus and major rivers are displayed (blue/teal, with names in black bold). Health zones comprising the focus are colour-shaded as follows: Angumu (pink), Logo (light pink/peach), Mahagi (purple/lilac), Nyarambe (light blue), Rethy (brown), with health areas outlined (thin black contours) with their names labels (haloed in white). The map was produced in ArcGIS Pro (Esri, Redlands, CA, USA) using publicly available spatial datasets (GRID3 COD – Health Areas v5.0 [27] and GRID3 COD – Health Zones v5.0 [28]). River and stream networks were derived from OpenStreetMap [35] and adjusted using ground-truth waypoints.

Current land cover on the highlands is predominantly savannah with fragmented primary and secondary forest blocks/patches, with forest more extensive to the west and a forest-savannah mosaic extending eastward [25]. The climate is humid tropical with weakly bimodal seasons (with some variation consisting of the main rains around July–October, main dry season November–March, minor rains in April–May and minor dry season in June) [17, 22] and approximately 1,400 millimetres of mean annual precipitation [26].

### 2.3 Evidence synthesis

We conducted a comprehensive scoping evidence synthesis of onchocerciasis and *Simulium* vectors in the Kakoi-Koda focus from 1980 to 31 July 2025, informed by PRISMA-ScR guidance (PRISMA extension for scoping reviews, as detailed by Tricco *et al.* 2018 [29]). Sources included PubMed, Google (Scholar) and the World Health Organization’s Institutional Repository for Information Sharing (WHO IRIS) using combinations of the following search terms: “onchocerciasis”, “*Simulium”*, “blackfly”, “Kakoi”, “Koda” and “Ituri” and their French equivalents (“onchocercose”, “simulie(s)” and “mouche noire”) in titles, abstracts or mentions in the text. The use of “Ituri” as a required term ensured the retrieval of studies from both before and after the area’s official designation as Ituri Province (formerly known as Ituri District within Orientale Province). In addition to electronic searches, we reviewed: (1) programme documents from the DRC National Onchocerciasis Elimination Programme, (2) data from the Expanded Special Project for Elimination of Neglected Tropical Diseases (ESPEN) portal for DRC [20], (3) unpublished reports and protocols generated within the NSETHIO research programme [30-32], and (4) reference lists from relevant review articles for additional sources. This approach allowed us to capture both peer-reviewed publications and grey-literature (reports) relevant to the context of the Kakoi-Koda focus.

The inclusion criteria were comprised of studies or reports with information on: (1) programmatic data (i.e., CDTI distribution and coverage), (2) onchocerciasis epidemiology (i.e., nodule palpation, anti-Ov16 serological and skin-snip surveys), and/or (3) entomology (i.e., human landing catches, blackfly testing for *O. volvulus* infection/infectivity, and breeding site prospections) from the Kakoi-Koda focus (i.e., data pertaining to at least one of its five HZs as illustrated in Fig 1). Exclusion criteria comprised studies and documents without geographical relevance to the focus or lacking extractable outcomes. Titles, abstracts and study reports were screened, and relevant data items were extracted (location, year, population, numbers examined, diagnostic method, key epidemiological/entomological/programmatic indicators, vector species) by LJA, summarised in Results subsection 3.1, and the full extraction provided in the (Supplementary) S1 Dataset. Discrepancies were resolved by discussion with the remaining co-authors.

### 2.4 Geospatial analysis of land cover and deforestation

We quantified forest cover change using the Global Forest Change dataset by Hansen *et al.* (30-metre resolution of per cent tree cover in 2000 and annual tree loss from 2001 to 2024) [33, 34]. Administrative boundaries for HZs and HAs in the focus were obtained from GRID3 COD – Health Areas v5.0 [27] and GRID3 COD – Health Zones v5.0 [28], respectively. Rivers and streams networks were extracted from OpenStreetMap to visualise waterways in the study area [35] and corrected based on ground-truth waypoints.

For the present geospatial analyses, the Hansen rasters were spatially restricted to the five HZs of the Kakoi-Koda focus and all layers were re-projected to a common coordinate system (WGS 1984 UTM Zone 36N). Forest in 2000 was defined as pixels with tree cover ≥30% (as used by Hansen *et al*. [33] and the definition followed by the DRC [36]), and forest loss as pixels that were forest in 2000 and had a non-zero loss year (2001–2024). Using zonal tabulations, we calculated for each HA the area of forest in 2000 and the area of that forest that was subsequently lost, and expressed deforestation as the percentage of 2000 forest cover lost by 2024. These HA-level estimates were then mapped, as well as summarised by HZ. We repeated this analysis with the definition of “dense forests” as pixels with tree cover ≥70% [37] to better capture closed-canopy habitats likely to be relevant for *Simulium neavei* [38]. Spatial analyses and map visualisations were performed in ArcGIS Pro (Esri, Redlands, CA, USA) and ArcGIS Online.

### 2.5 Interactive web map and data availability

To complement the static figures, we assembled an interactive web map (ArcGIS Online) that overlays, by survey year and data type, the epidemiological and entomological observations together with geospatial context. In parallel, all epidemiological, entomological, programmatic and geospatial inputs (including previously unpublished NSETHIO surveys and reports) were harmonised into a single master dataset accompanying this article (S1 Dataset). The dataset includes standardised variables, harmonised geographical identifiers, and documentation of data provenance and processing steps.

The web map includes: 1) REMO nodule surveys; 2) anti-Ov16 serological surveys (children; adults; combined); 3) skin-snip microscopy surveys (population; adolescents only; adolescents and adults); 4) freshwater-crab trapping data for *S. neavei*-associated breeding; 5) breeding-site inspections for blackfly larvae; and 6) human-biting blackfly data using human landing catch (HLC). Additionally, map layers include health-zone/health-area boundaries, rivers/streams, and annual tree-cover loss since 2000. Each layer has metadata describing source, dates, methods and any harmonisation applied. The ArcGIS interactive item is publicly accessible online (https://arcg.is/1PDyO0), and a README file is available in (Supplementary) S1 Appendix Text A. To protect privacy, all data are aggregated at the village level and contain no personal identifiers.

### 2.6 Statistical analysis

We analysed data on programmatic treatment coverage, *O. volvulus* epidemiology (REMO nodule prevalence categories, anti-Ov16 seropositivity and skin snip (microfilarial) prevalence) and entomology (simuliid larval breeding site prospections, and HLC data). Non-normally distributed (non-Gaussian) continuous variables were summarised as medians with interquartile ranges (IQRs, 1st quartile to 3rd quartile). Categorical data are presented as counts and percentages, and, where relevant, 95% confidence intervals (95% CIs) were computed using the Wilson score method with continuity correction [39, 40]. Because study designs, diagnostics, age groups and protocols varied across data sources, we harmonised metrics by data type and protocol, year data were collected, GPS coordinates (i.e., by HA and HZ for epidemiology data; river basins for entomology; village level for the interactive web map) and age strata (epidemiology: all ages ≥3 years for population prevalence data; children and adolescents as indicators of recent transmission). GPS coordinates were converted into decimal degrees and mapped in ArcGIS.

To characterise age- and sex-specific cumulative exposure to *O. volvulus*, we used data from the cross-sectional community anti-Ov16 rapid diagnostic test (RDT) survey in Logo HZ in 2016, identified in the evidence synthesis, restricted to residents aged ≥3 years with valid serology (n = 921). We focused on this community-based dataset because the remaining serology datasets were not population-representative. Ages were grouped into 3–6, 7–10, 11–20, 21–30, 31–40, 41–50 and ≥51 years to ensure sufficient sample sizes per band. For each age group and sex, we estimated anti-Ov16 seroprevalence with 95% CIs using the Wilson score method with continuity correction and displayed these estimates in an age- and sex- specific seroprevalence plot. We fitted a logistic regression model with age, sex and their interaction, adjusting for health area, to assess age and sex differences in cumulative exposure.

For person-level paired comparisons of skin-snip microscopy and anti-Ov16 RDT conducted in the field in children aged 3–10 years (indicative of ongoing transmission), we reported per cent agreement and Cohen’s kappa (κ) with asymptotic 95% CIs. Marginal homogeneity was assessed using McNemar’s test with mid-p two-sided p-values due to the small number of discordant pairs [41]. Skin-snip results were taken as a reference to estimate the sensitivity and specificity of anti-Ov16 RDT in the field with exact (Clopper-Pearson) 95% CIs, given the small sample size and sparse discordant data. Analyses were performed in R (version 4.4.1) using base functions and the packages DescTools (κ, CIs) [42] and epiR (diagnostic accuracy) [43].

## 3. RESULTS

### 3.1 Evidence base and data coverage

We assembled epidemiological, entomological and control programme data for the Kakoi-Koda focus from 1980 to 2025. Table 1 summarises designs, populations, procedures, metrics, years, areas and key limitations for each data stream. Epidemiological sources included REMO nodule palpation, anti-Ov16 serology from community, case-control and cohort surveys investigating OAE, and community and study-based skin-snip surveys (including screening datasets from the 2009–2011 and 2021–2023 moxidectin clinical trials). Entomology encompassed targeted inspections of breeding sites and freshwater crabs, (informal) “spot checks” of human-biting blackfly species by HLC and use of genus-specific mitochondrial 16S rDNA gene and species-specific NADH dehydrogenase subunit 5 (ND5) gene polymerase chain reaction (PCR) for the detection of *O. volvulus* DNA in flies’ heads (infectivity prevalence) or heads and bodies (infection prevalence) in blackfly samples. Programme documents provided contextual information on ivermectin distribution history (i.e., geographical and therapeutic coverage) by HZ and calendar year. The next subsections detail (3.1.1) CDTI delivery histories by HZ and HA, (3.1.2) longitudinal trends in infection indicators, and (3.1.3) vector species and transmission ecology.

**Table 1.**
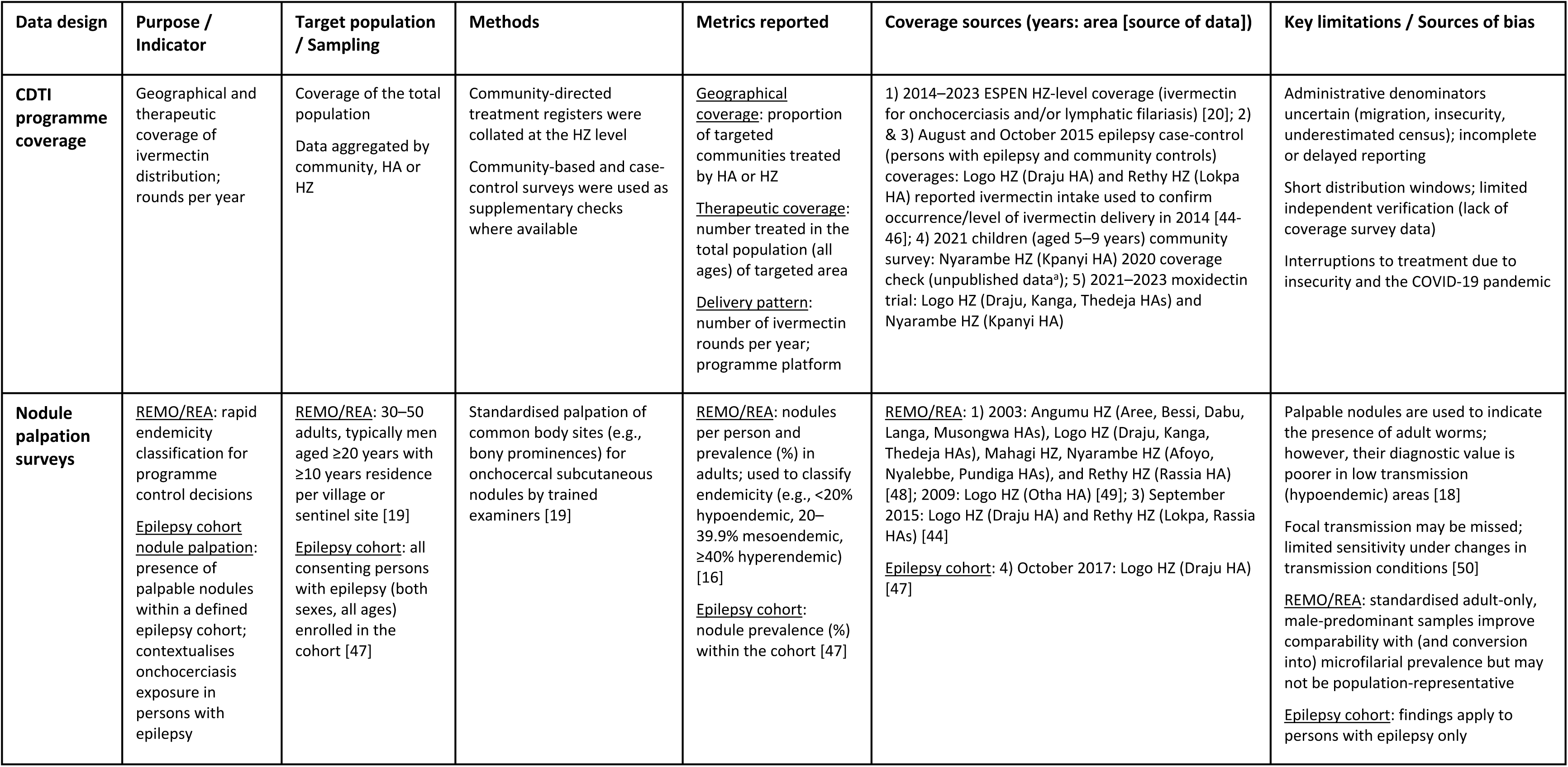
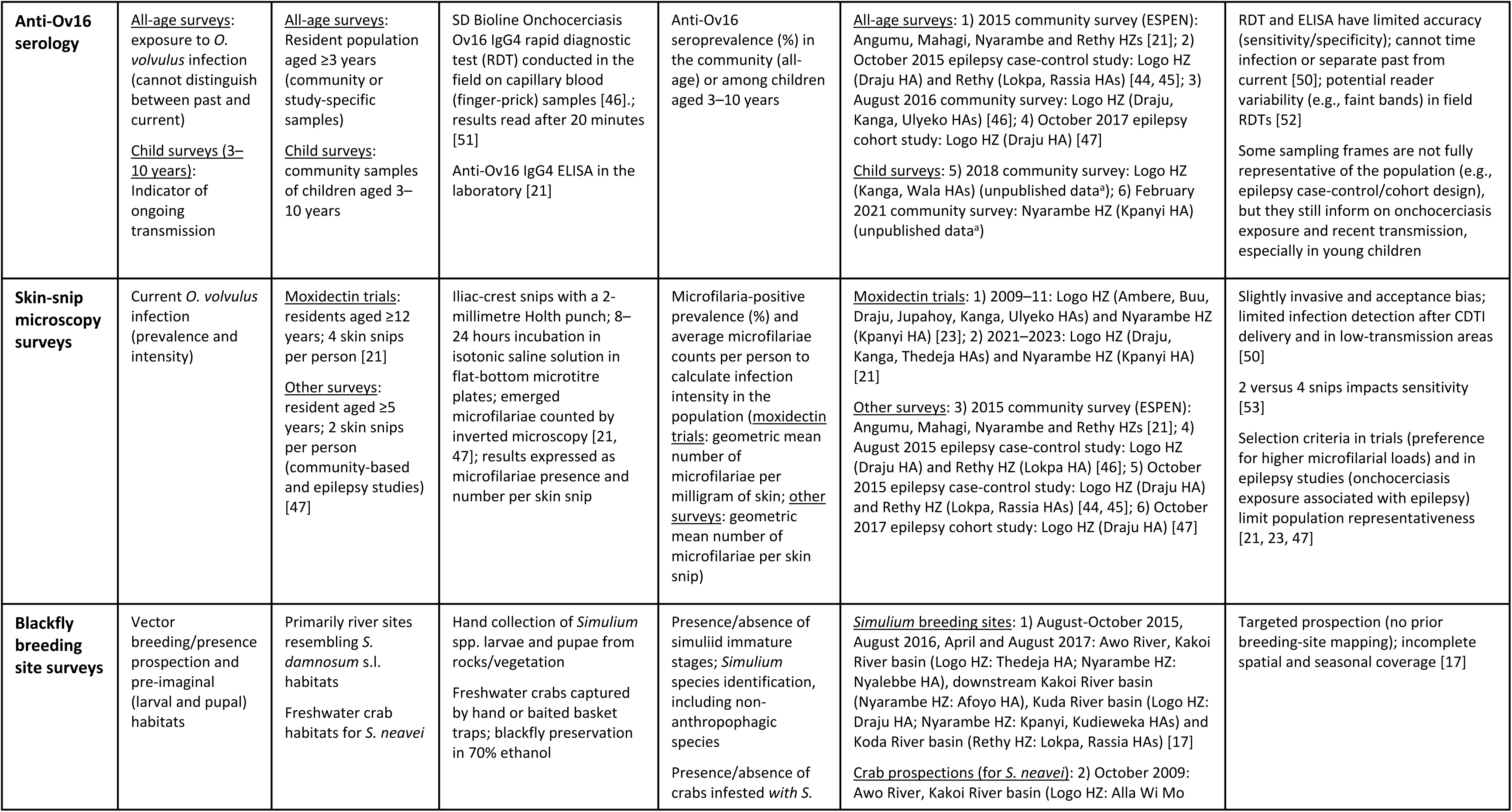
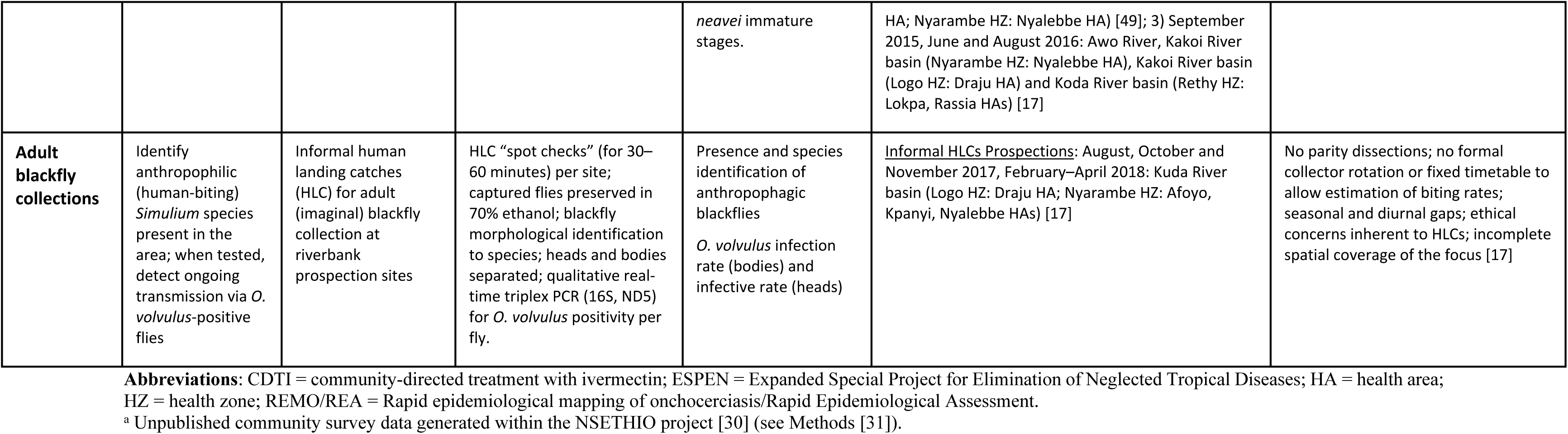
Epidemiological and entomological data sources and methods in the Kakoi–Koda onchocerciasis focus, Ituri Province, DRC.

#### 3.1.1 CDTI delivery histories by health zone (HZ) and health area (HA)

Ivermectin delivery in the Kakoi-Koda focus differed between HZs and by target disease. For onchocerciasis control, CDTI began in Angumu HZ in 2009 (ESPEN repository-derived therapeutic coverages of 80–82% of the total population) and in Rethy HZ in 2012 (68–81% coverage). In contrast, in Nyarambe HZ, mass drug administration with ivermectin and albendazole for lymphatic filariasis commenced in 2016 (80–84% coverage) [20]. In Logo and Mahagi HZs, CDTI had not been implemented as of 2025 [20]. Ivermectin delivery was temporarily interrupted in Ituri in 2020 due to the COVID-19 pandemic [20].

Beyond routine CDTI, clinical trials evaluating microfilaricidal treatments have taken place in Logo and Nyarambe HZs (only treating recruited participants). Firstly, in 2009–2011, the first moxidectin trial (Phase III 3110A1-3000) administered single doses to 315 (8 milligrams of moxidectin) and 157 (150 micrograms of ivermectin/kilogram of body weight) participants in Logo HZ (Draju and Kanga HAs) [24, 54]. Secondly, 197 persons living with epilepsy in Logo HZ (Draju, Kanga, Wala, Ulyeko, Thedeja HAs) received one to three ivermectin doses during 2017–2018 in a proof-of-concept study [55, 56]. Only 1.5% (3/197) of participants in this second trial self-reported prior ivermectin use [55]. Thirdly, the second moxidectin trial in the DRC (Phase IIIb MDGH-MOX-3002) enrolled additional participants in 2021–2023 who received either moxidectin or ivermectin (trial results pending) [21, 57]. The latter trial recorded self-reported prior ivermectin use in the preceding five years in 0.4% (29/7,576) of screened individuals from Logo HZ (Logo, Kanga, Thedeja HAs) and 0.9% (9/1,056) from Nyarambe HZ (Kanga HA) [58]. Finally, an ongoing trial evaluating the safety and efficacy of multiple annual or biannual doses of moxidectin and ivermectin began in 2021 and is ongoing until 2026 (Phase IIIb MDGH-MOX-3001, results pending) [21, 59].

In addition to the clinical trials, two epilepsy case-control surveys provided local verification of ivermectin intake. The first survey was conducted around August 2015, with a population sampled aged 3–75 years (median: 22 years, IQR: 14–30 years) and 52.4% females. The second survey was conducted in October 2015, with a population sampled aged 4–35 years (median: 17 years, IQR: 12–24 years) and 53.6% females. The former survey (August) found a self-reported ivermectin intake in 2014 of 1.5% (1/69) in Logo HZ (Draju HA) and of 75.4% (43/58) in Rethy HZ (Kpandroma, Lokpa HAs) [44, 45]. The latter survey (October) found a self-reported ivermectin intake in 2014 of 9.1% (4/45) in Logo HZ (Draju HA) and of 73.3% (11/15) in Rethy HZ (Lokpa HA) [44, 45]. Among respondents who reported taking ivermectin in the latter survey, the median cumulative number of ivermectin rounds taken was 3 (IQR: 3–3; range: 0–3 rounds).

An unpublished, self-reported community survey conducted in 2020 in Nyarambe HZ (Kanga HA) among ivermectin-eligible children aged 5–9 years recorded a therapeutic coverage of 91.6% (87/95). Among treated children, the median cumulative number of ivermectin rounds received by 2020 was 2 (IQR: 2–2; range: 1–3 rounds). Participants had a median age of 7 years (IQR: 7–9 years), and 56.4% were female.

#### 3.1.2 Longitudinal trends in *O. volvulus* infection indicators

Across data sources, indicators of *O. volvulus* infection declined between early observations (REMO 2003; skin-snip screening data in 2009–2011) and more recent surveys (in 2015–2023). To document this pattern, we first summarise baseline endemicity from nodule palpation surveys, then present serological evidence (all-age exposure and child anti-Ov16 as an indicator of recent transmission), and finally describe skin-snip prevalence and intensity between the 2009–2011 and 2021–2023 moxidectin trial and OAE studies (2015–2017).

##### Nodule palpation prevalence to establish baseline endemicity

A proportion of adult *O. volvulus* worms reside in subcutaneous nodules or onchocercomata [60], making nodule palpation a useful REA method for the evaluation of prevalence in areas of moderate to high endemicity. In 2003, REMO/REA nodule palpation identified several areas as meso- or hyperendemic (≥20% nodule prevalence) for onchocerciasis, permitting an initial delineation of the Kakoi-Koda focus (Fig 2) [48]. A median of 30 individuals (IQR: 30–40) aged ≥20 years per HA (one village) were examined, of which 81.8% (637/779) were male.

Hyperendemicity (nodule prevalence ≥40%) appeared concentrated between the Awo River (northeast Kuda branch) and the Kakoi, Kuda and Lebu River basins, in the following HZs: 1) Angumu: Bessi (83.3%, 95% CI: 64.5–93.7), Dabu (90.0%, 95% CI: 72.3–97.4) and Langa HAs (70.0%, 95% CI: 50.4–84.6); 2) Logo: Draju (97.6%, 95% CI: 85.6–99.9) and Kanga HAs (70.0%, 95% CI: 50.4–84.6); and 3) Nyarambe: Afoyo HA (52.0%, 95% CI: 37.6–66.1).

Mesoendemicity (nodule prevalence ≥20% but <40%) extended upstream in the Kakoi and the Koda and Loda River basins: 1) Angumu: Aree (36.0%, 95% CI: 23.3–50.9) and Musongwa HAs (34.7%, 95% CI: 22.1–49.7); 2) Nyarambe: Pundiga HA (30.0%, 95% CI: 15.4–49.6); 3) Rethy: Rassia HA (36.7%, 95% CI: 20.5–56.1). Hypoendemic areas (nodule prevalence ≥5% but <20%) were recorded around the Awo River: 1) Logo: Thedeja HA (6.7%, 95% CI: 1.2–23.5); 2) Nyarambe: Nyalebbe HA (16.0%, 7.6–29.7). An additional survey in 2009 in Otha HA (Logo HZ) classified the area as non-endemic (1/25, 4.0%, 95% CI: 2.1 –22.3) [49]. (Areas with <10% microfilarial prevalence are considered non-endemic/sporadic endemicity, corresponding to a nodule prevalence <5%[15, 61, 62].)

Using similar methods, a 2015 REA study was conducted in several villages within three HAs that reported high prevalence of onchocerciasis-associated epilepsy: Draju HA (26 individuals screened) in Logo HZ; and Lokpa (29) and Rassia (54) HAs in Rehty HZ [44]. Nodule prevalence was 34.6% (95% CI: 17.9–55.6) in Draju, 29.6% (95% CI: 18.4–43.8) in Rassia, and 17.2% (95% CI: 6.5–36.5) in Lokpa. Among those with nodules, the median number was one (IQR: 1–1; range: 1–6). A last nodule palpation survey in October 2017 examined 38 adults aged ≥20 years living with epilepsy in Draju HA (Logo HZ), including the same villages as in the 2003 and 2015 surveys, and recorded a nodule prevalence of 21.1% (95% CI: 10.1–37.8) [47].

##### All-age anti-Ov16 serological surveys to assess *O. volvulus* cumulative exposure

Anti-Ov16 serological tests were performed (with whole blood) using the SD Bioline anti-Ov16 RDT (Onchocerciasis IgG4 rapid test, Abbott Standard Diagnostics, Inc., Yongin, Korea; method detailed in Table 1). All-age (≥3 years) anti-Ov16 RDTs serosurveys conducted during 2015–2017 indicate high cumulative exposure to *O. volvulus* in both Logo and Rethy HZs (Table 2), in agreement with 2003 REMO classifications in Fig 2. Seropositivity was consistently higher in Draju HA (Logo HZ). In the 2015 epilepsy case-control sampling, seroprevalence in Draju was 41.2% (95% CI: 32.2–50.8), closely followed by Rassia HA (Rethy HZ) at 37.8% (95% CI: 22.9–55.2), and lower in Lokpa HA (Rethy HZ) at 29.4% (95% CI: 17.9–44.0) [44, 45]. These patterns are consistent with the 2015 REA described above, with moderate nodule prevalence in Draju and Rassia (30–35%) and lower prevalence in Lokpa (17%).

**Table 2.** All-age (≥3 years) anti-Ov16 seroprevalence by health zone and health area (2015–2017), measured by field rapid diagnostic test.

| Health Area | Study number [source of data] | Year | Study design; population | Median age (IQR; range) years | Proportion of females % | Anti-Ov16 seropositive/total tested | Anti-Ov16 seroprevalence % (95% CI) |
| --- | --- | --- | --- | --- | --- | --- | --- |
| **Logo Health Zone** |  |  |  |  |  |  |  |
| Draju | 1 [44, 45] | 2015 | Case-control; 56 epilepsy cases and 58 controls | 17 (12–25; 4–60) | 51.1 | 47/114 | 41.2 (32.2–50.8) |
|  | 2 [46] | 2016 | Cross-sectional; 438 community samples | 16 (9–35; 3–88) | 55.0 | 158/438 | 36.1 (31.6–40.8) |
|  | 3 [47] | 2017 | Epilepsy cohort; 73 samples | 21 (16–28; 3–59) | 53.4 | 47/73 | 64.4 (52.2–75.0) |
| Kanga | 2 [46] | 2016 | Cross-sectional; 476 community samples | 16 (8–34; 3–85) | 51.1 | 123/476 | 25.8 (22.0–30.1) |
| Ulyeko | 2 [46] | 2016 | Cross-sectional; 7 community samples | 28 (13–46; 3–48) | 28.6 | 2/7 | —<sup>c</sup> |
| Mixed <sup>a</sup> | 3 [47] | 2017 | Epilepsy cohort; 286 samples | 20 (14–30; 3–72) | 48.3 | 80/286 | 28.0 (22.9–33.6) |
| **Rethy Health Zone** |  |  |  |  |  |  |  |
| Lokpa | 1 [44, 45] | 2015 | Case-control; 24 epilepsy cases and 27 controls | 17 (12–28; 5–64) | 66.7 | 15/51 | 29.4 (17.9–44.0) |
| Rassia | 1 [44, 45] | 2015 | Case-control; 21 epilepsy cases and 16 controls | 15 (12–21; 7–55) | 31.6 | 14/37 | 37.8 (22.9–55.2) |

**Abbreviations:** 95% CI = 95% confidence interval; IQR = Interquartile range (1<sup>st</sup> quartile to 3<sup>rd</sup> quartile).
<sup>a</sup> Most participants in the 2017 epilepsy cohort in Logo Health Zone (Kanga, Ulyeko, Thedeja and Wala Health Areas) lacked village information, so individual records could not be assigned to a specific health area.
<sup>b</sup> Study number is the numerical label (1–3) grouping rows belonging to the same data collection round; identical numbers indicate the same study conducted in multiple health areas.
<sup>c</sup> Proportion and 95% confidence interval not reported for very small samples (number < 10).
Note: Direct statistical comparisons between studies should be avoided because study designs, sampling frames and age structures differ.

In the 2016 community sampling, Draju seroprevalence remained elevated at 36.1% (95% CI: 31.6–40.8) compared with 25.8% (95% CI: 22.0–30.1) in Kanga HA (Logo HZ) [46]. The estimate for Ulyeko HA (Logo HZ) is based on a very small sample (2/7 seropositive), providing limited information beyond indicating some *O. volvulus* exposure in that HA. In the 2017 epilepsy cohort, a seroprevalence of 64.4% (95% CI: 52.2–75.0) was documented in Draju, statistically significantly higher than the 28.0% (95% CI: 22.9–33.6) seroprevalence from the pooled Kanga/Ulyeko/Thedeja/Wala HAs (Logo HZ) cohort [47]. Given that this cohort comprised persons with epilepsy, the majority fulfilling the OAE criteria [47], a higher seroprevalence compared with community-based surveys is expected. Accordingly, because study designs and age structures differed (i.e., case-control versus community versus cohort), no formal between-study statistical comparisons were undertaken.

Age- and sex-specific anti-Ov16 seroprevalence from the 2016 community samples survey in Logo HZ (Table 2) is shown in Fig 3. Seroprevalence rose steeply from childhood to early adulthood and then plateaued around 60–70% in older adults, consistent with the hyperendemic status indicated by the 2003 REMO survey. Seroprevalence tends to be higher in males in the younger age groups, but higher in females in the older age groups. In a logistic regression including a sex-by-age-group interaction and adjusting for HA, there was no strong evidence of sex differences up to 40 years of age (odds ratio female versus male (OR_f/m_): 0.77, p = 0.19 for ages 3–30 years; OR_f/m_: 0.92, p = 0.85 for 31–40 years), although point estimates suggested slightly higher exposure in young men. In contrast, among adults aged >40 years, women had markedly higher seroprevalence than men (OR_f/m_: 2.46, p = 0.004).

**Fig 3.**
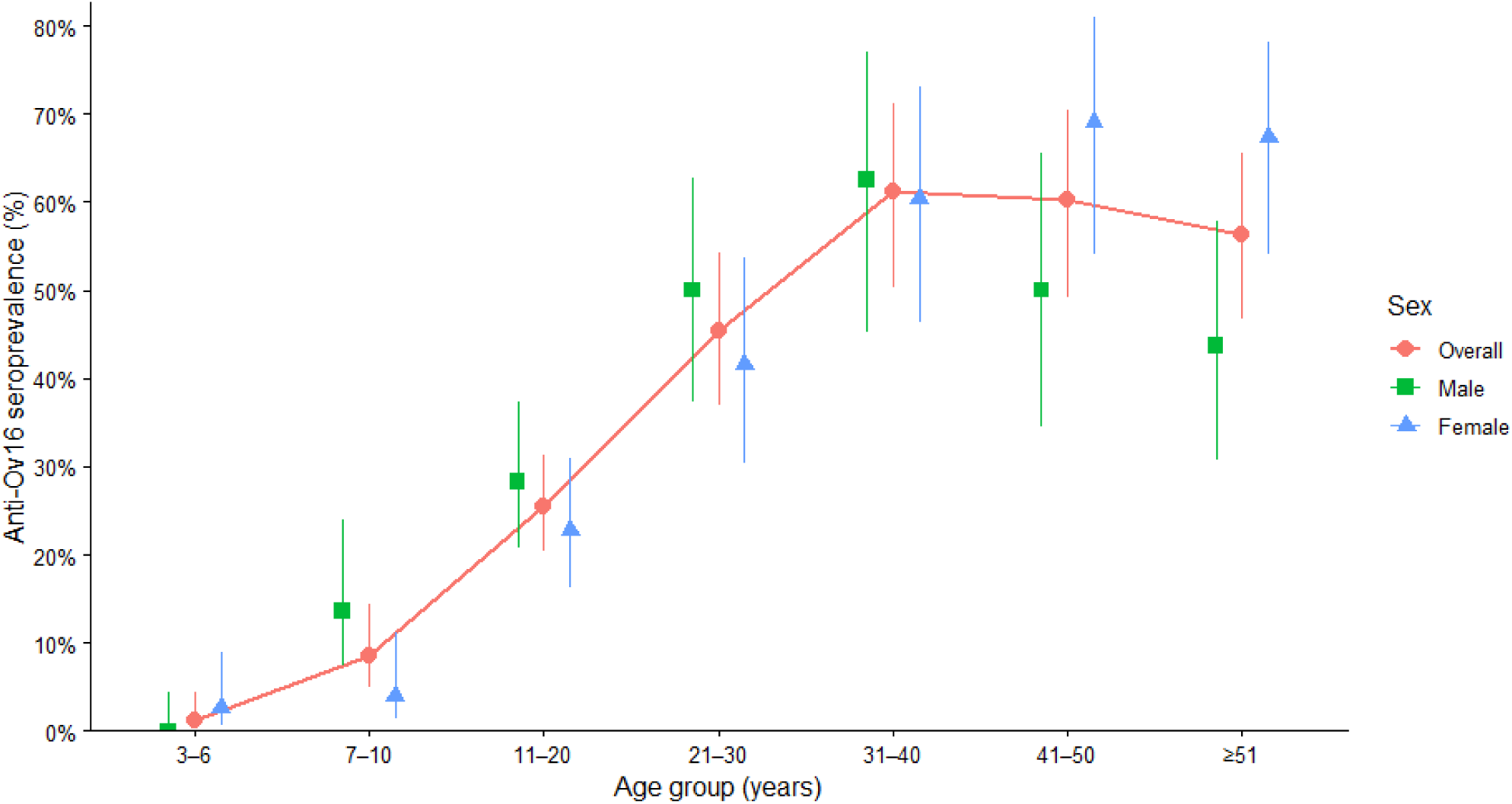
Age-specific anti-Ov16 seroprevalence by sex in the 2016 community survey in Logo Health Zone (Draju, Kanga and Ulyeko Health Areas) [46]. Red circles (joined by red solid line) show seroprevalence estimates with 95% Wilson confidence intervals (error bars) by age group overall, with green squares indicating males and blue triangles indicating females (male and female estimates are slightly offset horizontally for better visualisation).

An ESPEN repository-derived anti-Ov16 ELISA dataset from 2015 targeted non-endemic villages surrounding the Kakoi-Koda focus, and therefore is presented in S1 Appendix Text B [21].

##### Child anti-Ov16 RDT serological surveys to indicate *O. volvulus* ongoing transmission

In Logo HZ (Draju HA), the 2015 epilepsy case-control survey found anti-Ov16 RDT seropositivity in children aged 3–6 (1/4) and 7–10 (2/12) years (Table 3) [44, 45]. In the subsequent 2016 community survey from the same locality, no seropositive children were detected among the 3–6-year group (0/70; 0%), whereas 10.3% of 7–10-year-olds were seropositive (7/68) [46]. A similar pattern was observed in Kanga HA (Logo HZ) in 2016, with seropositivity of 2.2% in 3–6-year-olds (2/90) and 9.6% in 7–10-year-olds (7/73). The epilepsy cohort study conducted across Draju, Kanga, Ulyeko, Thedeja, and Wala HAs (Logo HZ) in 2017 identified no seropositive children (0/30 in those aged 3–6 years; 0/26 in those aged 7–10 years) [47]. In 2018, a smaller, unpublished community survey in Logo HZ detected no seropositive 3–6-year-olds in Kanga (0/4) and 4.8% seropositivity among 7–10-year-olds (1/21). In Wala HA, no seropositive children were recorded (0/14 in those aged 3–6 years; 0/21 in those aged 7–10 years).

**Table 3.**
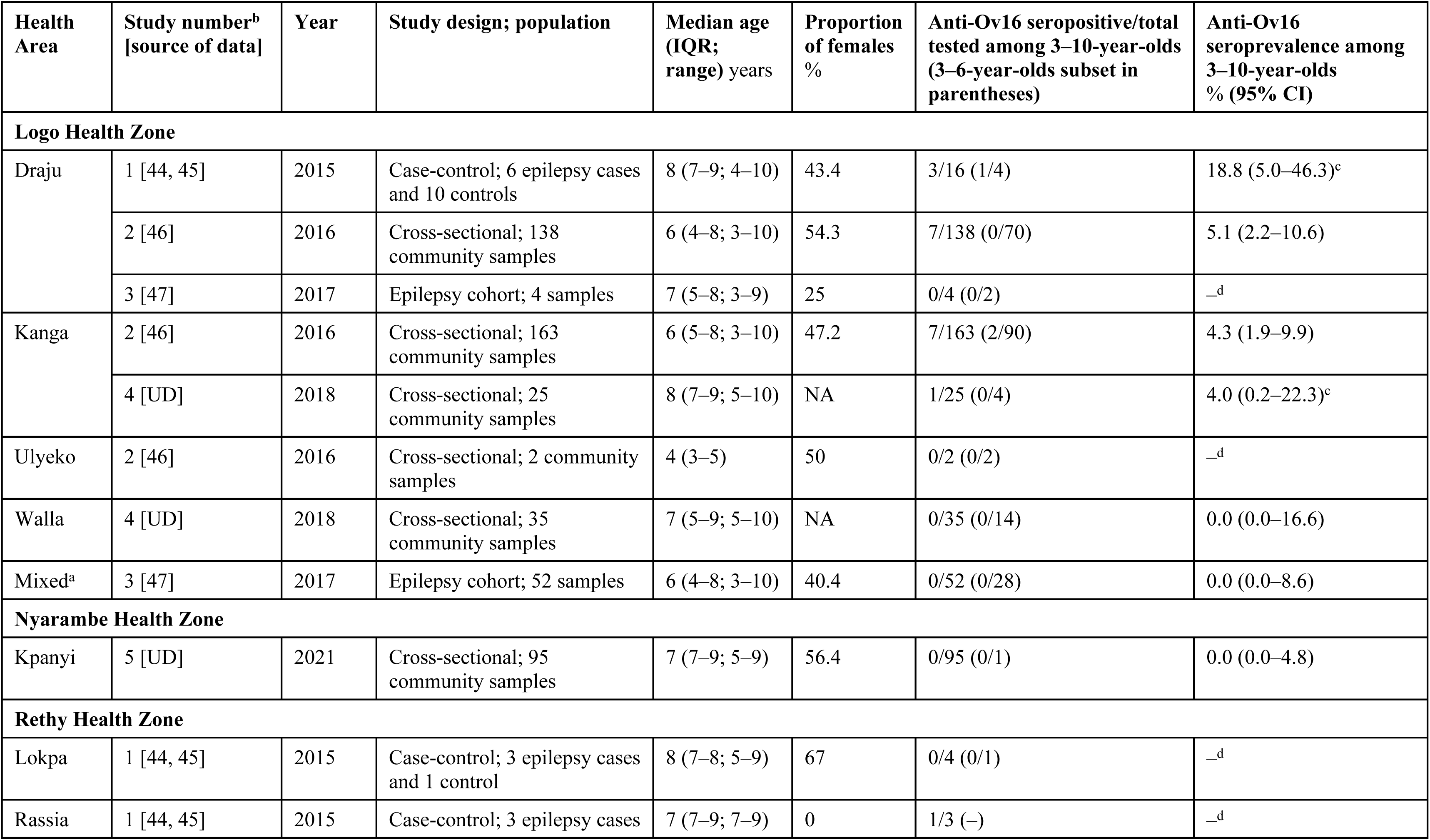
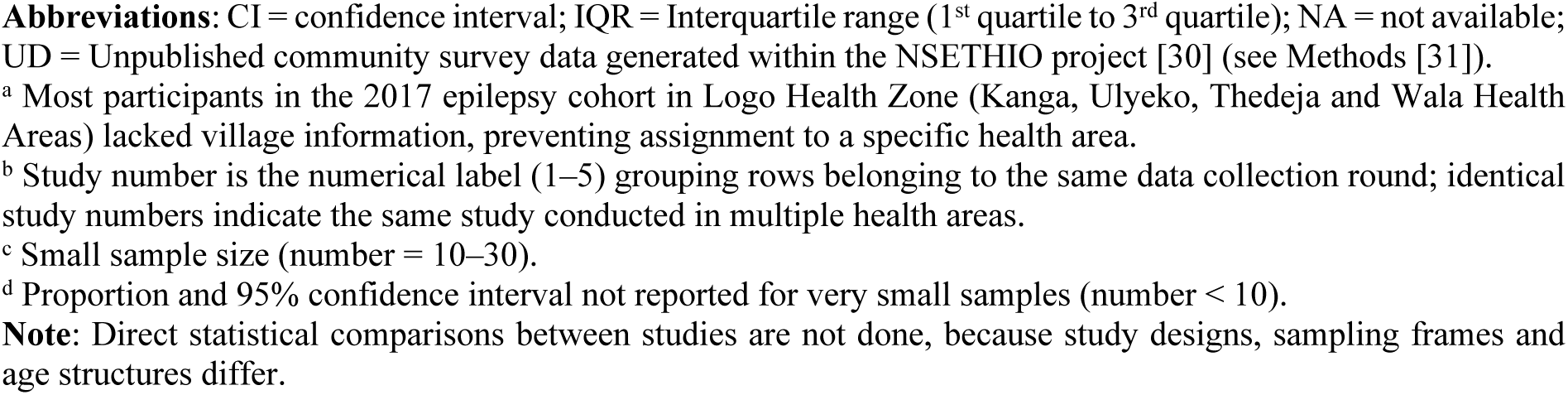
Child (aged 3–10 years) anti-Ov16 seroprevalence by health zone and health area (2015–2021), measured by field rapid diagnostic test. Anti-Ov16 seroprevalence is shown with Wilson score 95% confidence intervals.

| Health Area | Study number<sup>b</sup><br>[source of data] | Year | Study design; population | Median age (IQR; range) years | Proportion of females % | Anti-Ov16 seropositive/total tested among 3–10-year-olds (3–6-year-olds subset in parentheses) | Anti-Ov16 seroprevalence among 3–10-year-olds % (95% CI) |
| --- | --- | --- | --- | --- | --- | --- | --- |
| **Logo Health Zone** |  |  |  |  |  |  |  |
| Draju | 1 [44, 45] | 2015 | Case-control; 6 epilepsy cases and 10 controls | 8 (7–9; 4–10) | 43.4 | 3/16 (1/4) | 18.8 (5.0–46.3)<sup>c</sup> |
|  | 2 [46] | 2016 | Cross-sectional; 138 community samples | 6 (4–8; 3–10) | 54.3 | 7/138 (0/70) | 5.1 (2.2–10.6) |
|  | 3 [47] | 2017 | Epilepsy cohort; 4 samples | 7 (5–8; 3–9) | 25 | 0/4 (0/2) | —<sup>d</sup> |
| Kanga | 2 [46] | 2016 | Cross-sectional; 163 community samples | 6 (5–8; 3–10) | 47.2 | 7/163 (2/90) | 4.3 (1.9–9.9) |
|  | 4 [UD] | 2018 | Cross-sectional; 25 community samples | 8 (7–9; 5–10) | NA | 1/25 (0/4) | 4.0 (0.2–22.3)<sup>c</sup> |
| Ulyeko | 2 [46] | 2016 | Cross-sectional; 2 community samples | 4 (3–5) | 50 | 0/2 (0/2) | —<sup>d</sup> |
| Walla | 4 [UD] | 2018 | Cross-sectional; 35 community samples | 7 (5–9; 5–10) | NA | 0/35 (0/14) | 0.0 (0.0–16.6) |
| Mixed<sup>a</sup> | 3 [47] | 2017 | Epilepsy cohort; 52 samples | 6 (4–8; 3–10) | 40.4 | 0/52 (0/28) | 0.0 (0.0–8.6) |
| **Nyarambe Health Zone** |  |  |  |  |  |  |  |
| Kpanyi | 5 [UD] | 2021 | Cross-sectional; 95 community samples | 7 (7–9; 5–9) | 56.4 | 0/95 (0/1) | 0.0 (0.0–4.8) |
| **Rethy Health Zone** |  |  |  |  |  |  |  |
| Lokpa | 1 [44, 45] | 2015 | Case-control; 3 epilepsy cases and 1 control | 8 (7–8; 5–9) | 67 | 0/4 (0/1) | —<sup>d</sup> |
| Rassia | 1 [44, 45] | 2015 | Case-control; 3 epilepsy cases | 7 (7–9; 7–9) | 0 | 1/3 (—) | —<sup>d</sup> |

**Abbreviations:** CI = confidence interval; IQR = Interquartile range (1<sup>st</sup> quartile to 3<sup>rd</sup> quartile); NA = not available; UD = Unpublished community survey data generated within the NSETHIO project [30] (see Methods [31]).
<sup>a</sup> Most participants in the 2017 epilepsy cohort in Logo Health Zone (Kanga, Ulyeko, Thedeja and Wala Health Areas) lacked village information, preventing assignment to a specific health area.
<sup>b</sup> Study number is the numerical label (1–5) grouping rows belonging to the same data collection round; identical study numbers indicate the same study conducted in multiple health areas.
<sup>c</sup> Small sample size (number = 10–30).
<sup>d</sup> Proportion and 95% confidence interval not reported for very small samples (number < 10).
**Note:** Direct statistical comparisons between studies are not done, because study designs, sampling frames and age structures differ.

In Rethy HZ, an epilepsy case-control survey in Lokpa and Rassia HAs in 2015 detected one seropositive child among seven tested across Rassia and Lokpa HAs (Table 3) [44, 45]. In Nyarambe HZ (Kpanyi HA), the unpublished community survey in 2021 among 95 children aged 7–9 years recorded 0% anti-Ov16 seroprevalence.

##### Skin-snip microscopy surveys for moxidectin trials

Skin-snip microscopy data were assembled from sources using two different protocols: 1) moxidectin trial screenings (2009–2011 and 2021–2023), which obtained four snips per participant from community-recruited, self-reported ivermectin-naïve individuals aged ≥12 years; and 2) other surveys (2015–2017), which obtained two snips per participant in OAE case-control and cohort designs (methods detailed in Table 1). Because snip number and study design differ, prevalence and intensity are only compared within (but not between) these two groups.

In the 2009–2011 moxidectin trial screening, large numbers of residents were screened in Logo HZ, mostly in Draju (number = 1,149) and Kanga (n = 161) HAs (Table 4) [21]. Additionally, a smaller number of residents were screened in Nyarambe HZ (Kpanyi HA; n = 36). Village-level sample sizes were small and heterogeneous (median: 7, IQR: 3–25, range: 1–324 residents per village). *O. volvulus* microfilarial prevalence reached 79% in Logo (908/1,149) and 72% in Nyarambe, consistent with the abovementioned REMO (2003) hyperendemic classifications and all-age anti-Ov16 findings (2015–2017; Table 2).

**Table 4.**
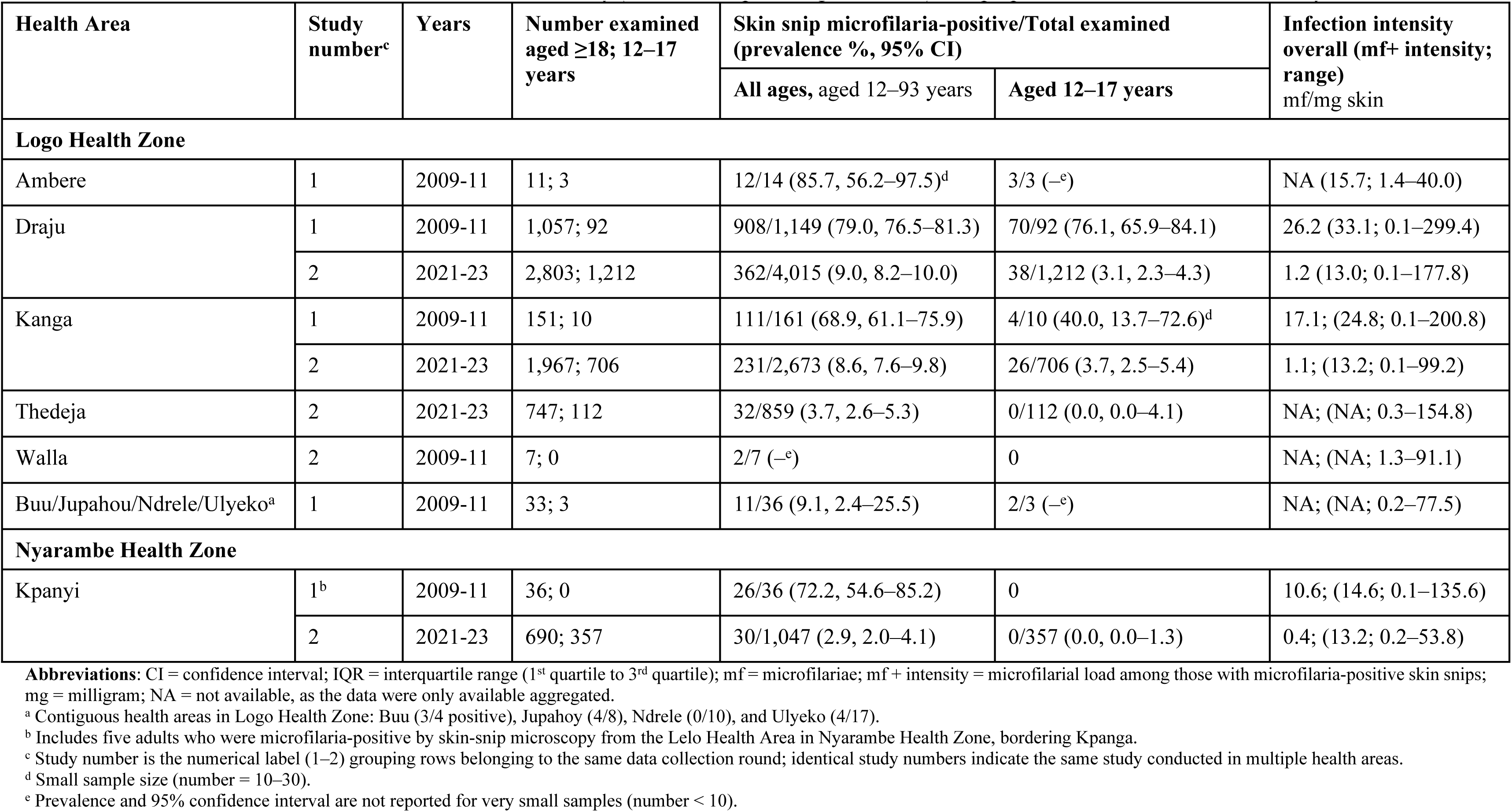
Skin-snip results from the moxidectin trials cross-sectional screenings in 2009–2011 and 2021–2023, by age group, health zone and health area. [21].

| Health Area | Study number<sup>c</sup> | Years | Number examined aged ≥18; 12–17 years | Skin snip microfilaria-positive/Total examined (prevalence %, 95% CI) |  | Infection intensity overall (mf+ intensity; range) mf/mg skin |
| --- | --- | --- | --- | --- | --- | --- |
|  |  |  |  | All ages, aged 12–93 years | Aged 12–17 years |  |
| Logo Health Zone |  |  |  |  |  |  |
| Ambere | 1 | 2009-11 | 11; 3 | 12/14 (85.7, 56.2–97.5)<sup>d</sup> | 3/3 (–<sup>e</sup>) | NA (15.7; 1.4–40.0) |
| Draju | 1 | 2009-11 | 1,057; 92 | 908/1,149 (79.0, 76.5–81.3) | 70/92 (76.1, 65.9–84.1) | 26.2 (33.1; 0.1–299.4) |
|  | 2 | 2021-23 | 2,803; 1,212 | 362/4,015 (9.0, 8.2–10.0) | 38/1,212 (3.1, 2.3–4.3) | 1.2 (13.0; 0.1–177.8) |
| Kanga | 1 | 2009-11 | 151; 10 | 111/161 (68.9, 61.1–75.9) | 4/10 (40.0, 13.7–72.6)<sup>d</sup> | 17.1; (24.8; 0.1–200.8) |
|  | 2 | 2021-23 | 1,967; 706 | 231/2,673 (8.6, 7.6–9.8) | 26/706 (3.7, 2.5–5.4) | 1.1; (13.2; 0.1–99.2) |
| Thedeja | 2 | 2021-23 | 747; 112 | 32/859 (3.7, 2.6–5.3) | 0/112 (0.0, 0.0–4.1) | NA; (NA; 0.3–154.8) |
| Walla | 2 | 2009-11 | 7; 0 | 2/7 (–<sup>e</sup>) | 0 | NA; (NA; 1.3–91.1) |
| Buu/Jupahou/Ndrele/Ulyeko<sup>a</sup> | 1 | 2009-11 | 33; 3 | 11/36 (9.1, 2.4–25.5) | 2/3 (–<sup>e</sup>) | NA; (NA; 0.2–77.5) |
| Nyarambe Health Zone |  |  |  |  |  |  |
| Kpanyi | 1<sup>b</sup> | 2009-11 | 36; 0 | 26/36 (72.2, 54.6–85.2) | 0 | 10.6; (14.6; 0.1–135.6) |
|  | 2 | 2021-23 | 690; 357 | 30/1,047 (2.9, 2.0–4.1) | 0/357 (0.0, 0.0–1.3) | 0.4; (13.2; 0.2–53.8) |

**Abbreviations:** CI = confidence interval; IQR = interquartile range (1<sup>st</sup> quartile to 3<sup>rd</sup> quartile); mf = microfilariae; mf + intensity = microfilarial load among those with microfilaria-positive skin snips; mg = milligram; NA = not available, as the data were only available aggregated.
<sup>a</sup> Contiguous health areas in Logo Health Zone: Buu (3/4 positive), Jupahoy (4/8), Ndrele (0/10), and Ulyeko (4/17).
<sup>b</sup> Includes five adults who were microfilaria-positive by skin-snip microscopy from the Lelo Health Area in Nyarambe Health Zone, bordering Kpanga.
<sup>c</sup> Study number is the numerical label (1–2) grouping rows belonging to the same data collection round; identical study numbers indicate the same study conducted in multiple health areas.
<sup>d</sup> Small sample size (number = 10–30).
<sup>e</sup> Prevalence and 95% confidence interval are not reported for very small samples (number < 10).

Community-recruited, self-reported ivermectin-naïve participants aged ≥12 years provided four skin snips each. *Onchocerca volvulus* infection prevalence is shown with Wilson score 95% confidence intervals, and infection intensity (microfilariae per milligram of skin). The proportion of females and males surveyed is not available.

Age-stratified skin snip results in Logo in 2009–2011 showed no clear signs of reduced infection in adolescents (Table 4). In particular, in Draju, overall prevalence was 79.0% (95% CI: 76.5–81.3), similar to the prevalence in 12–17-year-olds of 76.1% (95% CI: 65.9–84.1); in Kanga, overall prevalence was 68.9% (95% CI: 61.1–75.9) and prevalence among 12–17-year-olds was 40.0% (95% CI: 13.7–72.6), with the wide adolescent CI overlapping the adult estimate.

In the 2021–2023 moxidectin trial screening, sampling effort increased across the same HAs of Draju (n = 4,016), Kanga (n = 2,673) and Kpanyi (n = 1,047; Table 4), with larger village-level samples (median: 50, IQR: 21–218, range: 3–950 residents per village). Prevalence was markedly reduced in all three HAs (approximately 3–9% overall prevalence). Declines were most pronounced in younger age groups. For example, in Draju, the prevalence among 12–17-year-olds fell from 76.1% (95% CI: 65.9–84.1) in 2009–2011 to 3.1% (95% CI: 2.3–4.3) in 2021–2023. In Kpanyi, although the 2009–2011 sample was limited (n = 29 across the same villages between surveys), repeat adult sampling showed a reduction from 72.7% (16/22) to 2.0% (1/49) (Fisher’s exact p < 0.001). Thedeja HA (in Logo HZ) was also screened in 2021–2023 (but not in 2009–2011) as the trial expanded sampling to identify additional *O. volvulus*-infected residents for enrolment, and it showed the lowest microfilarial prevalence among the four HAs (3.7%, 95% CI: 2.6–5.3), consistent with its hypoendemic status according to the 2003 REMO surveys.

Infection intensity mirrored these prevalence declines (Table 4). Community infection intensity (i.e., microfilariae per milligram of skin) was substantially lower in 2021–2023 than in 2009–2011. Illustratively, Draju decreased from 26.2 (range: 0.1–299.4) to 1.2 (range: 0.1–177.8), Kanga from 17.1 (range: 0.1–200.8) to 1.1 (range: 0.1–99.2) and Kpanyi from 10.6 (range: 0.1–135.6) to 0.4 (range: 0.2–53.8). This decrease in microfilarial load was also observed when only considering those with microfilaria-positive skin snips (i.e., mf+ intensity in Table 4).

Across smaller 2009–2011 screening sites (Table 4), Ambere (12/14 positive) and Walla (2/7) HAs in Logo HZ showed microfilaria-positive skin-snip prevalence, consistent with ongoing transmission at that time and with REMO (2003) classifications of hyperendemicity in the contiguous Draju and Kanga HAs. The remaining samples in other HAs of Logo HZ suggested a northward decline in microfilarial prevalence from Draju (908/1,149) through Ulyeko (4/17) and Buu/Jupahoy (7/12 combined), with Ndrele (northernmost HA tested, 0/10) without microfilaria-positive skin snips (Fig 2 indicates HA locations and rivers). Given the very small denominators, these observations should be interpreted cautiously and not taken as a formal trend; nevertheless, they are congruent with REMO (2003) results, which classified Ukebu Ngali, the HA contiguous to Ndrele to the north, as non-endemic/sporadic endemicity.

##### Skin-snip microscopy surveys from other studies

Across the 2015–2017 epilepsy case-control and cohort surveys in Logo (e.g., Draju HA) and Rethy (e.g., Lokpa and Rassia HA) HZ, the highest all-age skin-snip prevalence values were consistently observed in Draju (range: 46–63%) compared to more moderate microfilaria-positive skin-snip prevalence values in other HAs (range: 20–47%; Table 5), concordant with the all-age anti-Ov16 seroprevalence for these areas (Table 2) [44-47]. Infection intensity was relatively high in most surveyed HAs (range: 13–93 microfilariae per skin snip), excluding Lokpa (range: 2–8 microfilariae per skin snip; Table 5), consistent with Lokpa’s hypo- to mesoendemicity status by nodule prevalence (2003–2015) compared to meso- to hyperendemicity in Rassia and Draju.

**Table 5.**
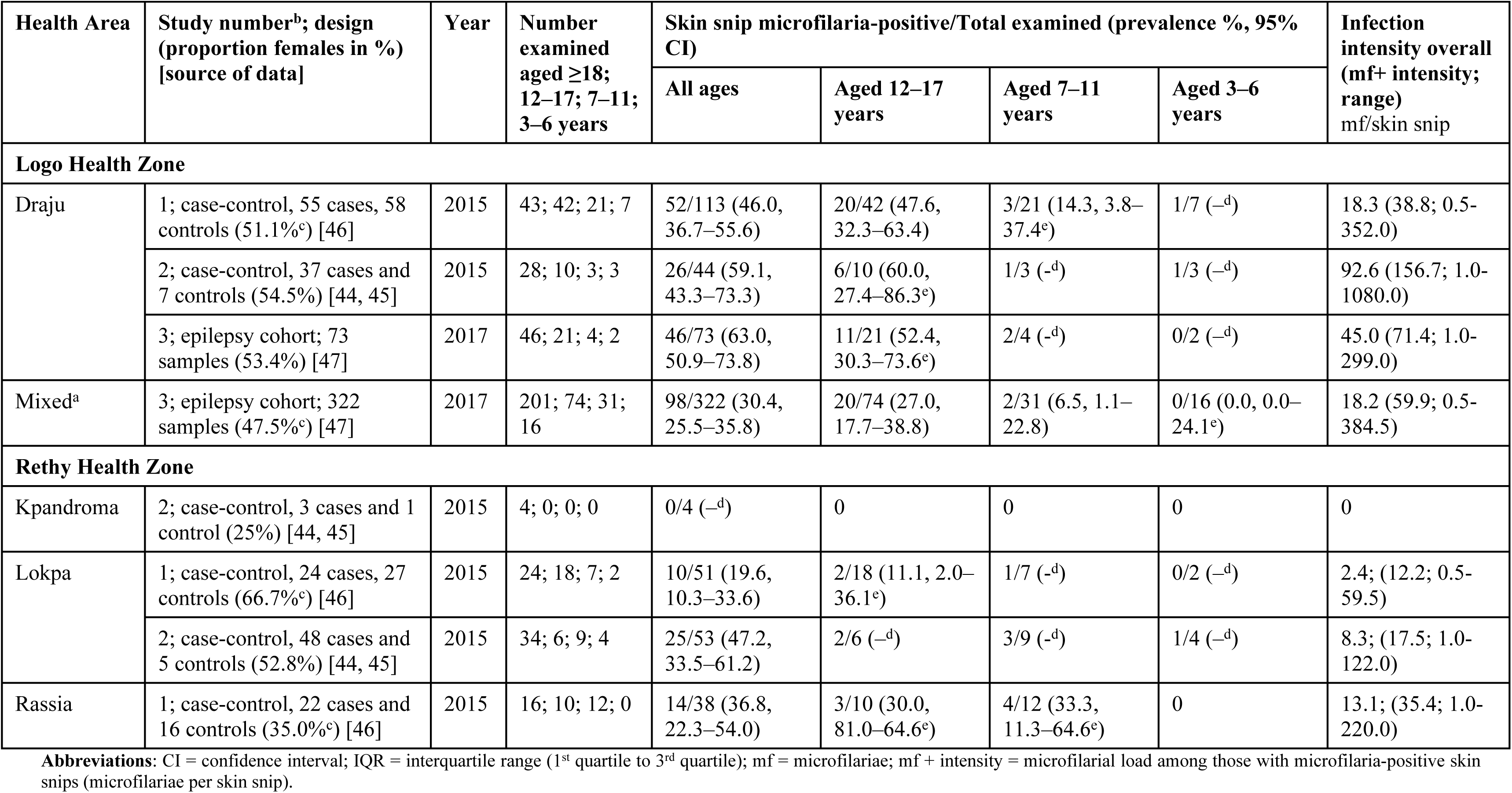
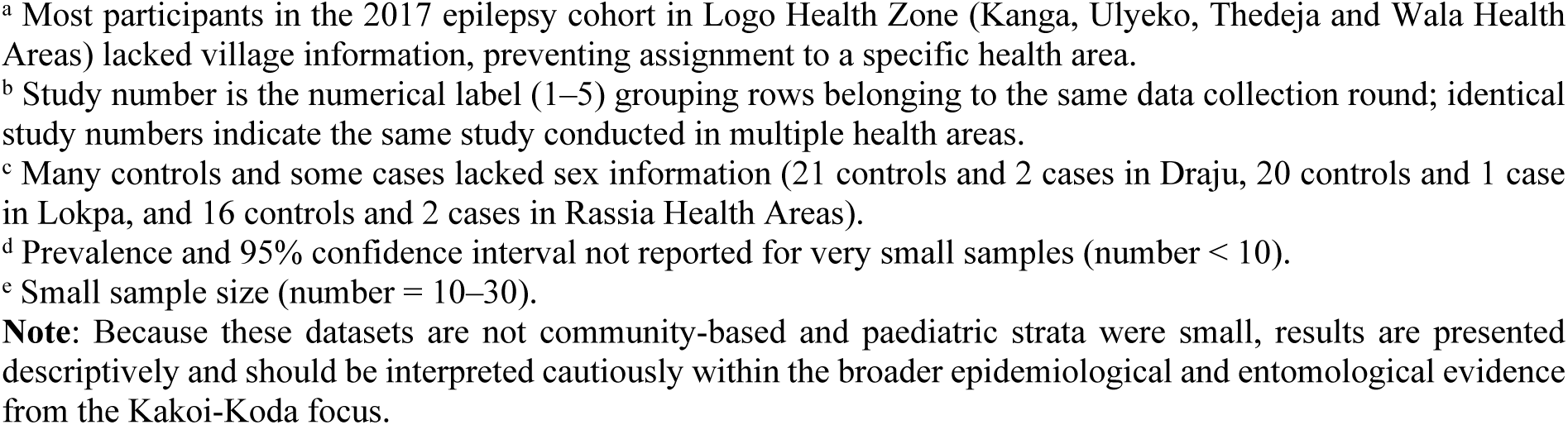
Skin-snip results from (other) studies conducted in 2015–2017 by age group, health zone and health area. Participants provided two skin snips each. *Onchocerca volvulus* microfilarial prevalence is shown with Wilson score 95% confidence intervals (CI), and infection intensity (microfilariae per skip snip). The proportion of females and males surveyed is not available.

| Health Area | Study number<sup>b</sup>; design<br>(proportion females in %)<br>[source of data] | Year | Number<br>examined<br>aged ≥18;<br>12–17; 7–11;<br>3–6 years | Skin snip microfilaria-positive/Total examined (prevalence %, 95%<br>CI) |  |  |  | Infection<br>intensity overall<br>(mf+ intensity;<br>range)<br>mf/skin snip |
| --- | --- | --- | --- | --- | --- | --- | --- | --- |
|  |  |  |  | All ages | Aged 12–17<br>years | Aged 7–11<br>years | Aged 3–6<br>years |  |
| Logo Health Zone |  |  |  |  |  |  |  |  |
| Draju | 1; case-control, 55 cases, 58 controls (51.1%<sup>c</sup>) [46] | 2015 | 43; 42; 21; 7 | 52/113 (46.0, 36.7–55.6) | 20/42 (47.6, 32.3–63.4) | 3/21 (14.3, 3.8–37.4<sup>e</sup>) | 1/7 (–<sup>d</sup>) | 18.3 (38.8; 0.5-352.0) |
|  | 2; case-control, 37 cases and 7 controls (54.5%) [44, 45] | 2015 | 28; 10; 3; 3 | 26/44 (59.1, 43.3–73.3) | 6/10 (60.0, 27.4–86.3<sup>e</sup>) | 1/3 (-<sup>d</sup>) | 1/3 (–<sup>d</sup>) | 92.6 (156.7; 1.0-1080.0) |
|  | 3; epilepsy cohort; 73 samples (53.4%) [47] | 2017 | 46; 21; 4; 2 | 46/73 (63.0, 50.9–73.8) | 11/21 (52.4, 30.3–73.6<sup>e</sup>) | 2/4 (-<sup>d</sup>) | 0/2 (–<sup>d</sup>) | 45.0 (71.4; 1.0-299.0) |
| Mixed<sup>a</sup> | 3; epilepsy cohort; 322 samples (47.5%<sup>c</sup>) [47] | 2017 | 201; 74; 31; 16 | 98/322 (30.4, 25.5–35.8) | 20/74 (27.0, 17.7–38.8) | 2/31 (6.5, 1.1–22.8) | 0/16 (0.0, 0.0–24.1<sup>e</sup>) | 18.2 (59.9; 0.5-384.5) |
| Rethy Health Zone |  |  |  |  |  |  |  |  |
| Kpandroma | 2; case-control, 3 cases and 1 control (25%) [44, 45] | 2015 | 4; 0; 0; 0 | 0/4 (–<sup>d</sup>) | 0 | 0 | 0 | 0 |
| Lokpa | 1; case-control, 24 cases, 27 controls (66.7%<sup>c</sup>) [46] | 2015 | 24; 18; 7; 2 | 10/51 (19.6, 10.3–33.6) | 2/18 (11.1, 2.0–36.1<sup>e</sup>) | 1/7 (-<sup>d</sup>) | 0/2 (–<sup>d</sup>) | 2.4; (12.2; 0.5-59.5) |
|  | 2; case-control, 48 cases and 5 controls (52.8%) [44, 45] | 2015 | 34; 6; 9; 4 | 25/53 (47.2, 33.5–61.2) | 2/6 (–<sup>d</sup>) | 3/9 (-<sup>d</sup>) | 1/4 (–<sup>d</sup>) | 8.3; (17.5; 1.0-122.0) |
| Rassia | 1; case-control, 22 cases and 16 controls (35.0%<sup>c</sup>) [46] | 2015 | 16; 10; 12; 0 | 14/38 (36.8, 22.3–54.0) | 3/10 (30.0, 81.0–64.6<sup>e</sup>) | 4/12 (33.3, 11.3–64.6<sup>e</sup>) | 0 | 13.1; (35.4; 1.0-220.0) |

**Abbreviations:** CI = confidence interval; IQR = interquartile range (1<sup>st</sup> quartile to 3<sup>rd</sup> quartile); mf = microfilariae; mf + intensity = microfilarial load among those with microfilaria-positive skin snips (microfilariae per skin snip).
<sup>a</sup> Most participants in the 2017 epilepsy cohort in Logo Health Zone (Kanga, Ulyeko, Thedeja and Wala Health Areas) lacked village information, preventing assignment to a specific health area.
<sup>b</sup> Study number is the numerical label (1–5) grouping rows belonging to the same data collection round; identical study numbers indicate the same study conducted in multiple health areas.
<sup>c</sup> Many controls and some cases lacked sex information (21 controls and 2 cases in Draju, 20 controls and 1 case in Lokpa, and 16 controls and 2 cases in Rassia Health Areas).
<sup>d</sup> Prevalence and 95% confidence interval not reported for very small samples (number < 10).
<sup>e</sup> Small sample size (number = 10–30).
**Note:** Because these datasets are not community-based and paediatric strata were small, results are presented descriptively and should be interpreted cautiously within the broader epidemiological and entomological evidence from the Kakoi-Koda focus.

Few microfilaria-positive skin snips were detected among children aged 3–11 years in Logo HZ (Table 5, 14–33%) compared with older age groups (48–60% in 12–17-year-olds; 46–63% in adults), in agreement with the low anti-Ov16 seroprevalence (by RDT) in Logo among 3–10-year-olds (0–19% in 2015–2018; Table 3). In contrast, child infection patterns in Rethy HZ did not show a similarly marked decline (33% in Rassia HA; Table 5) relative to adolescents (30% in Rassia among 12–17-year-olds) and adults (37% in Rassia).

An ESPEN repository-derived skin-snip dataset from 2015 sampled non-endemic villages surrounding the Kakoi-Koda focus, and therefore is presented in S1 Appendix Text B [21].

#### 3.1.3 Vector species and transmission ecology

To characterise potential vectors and where transmission may persist, we combined information from targeted larval/pupae breeding site prospections in both crab-associated and non-crab river habitats, together with informal “spot-check” HLCs to identify anthropophagic *Simulium* species and individual PCR testing of a subset for *O. volvulus* 16S and ND5 genes across the main basins of the focus (methods detailed in Table 1).

##### Breeding sites prospection

Crab-associated prospections for the *S. neavei* group conducted in October 2009 detected *S. neavei* larvae at one site within the focus, specifically at the Kakoi River basin (larvae in 11/12 crabs in Lelo HA, Nyarambe HZ) [49]. Two crabs from the south-eastern Awo River (Nyalebbe HA of Nyarambe HZ) were not infested with larvae, and no crabs were found further north-west at the limit of the focus (Alla Wi Mo HA, Logo HZ). In 2015–2016, no infested crabs were detected despite targeted sampling along the Kuda River basin (Logo HZ, Draju HA: 0/35 crabs from nine sites), the Koda River basin (Rethy HZ, Lokpa HA: 0/2 crabs from three sites; Rassia HA: 0/4 crabs from one site), and the Luda River basin (Lokpa HA: 0/30 crabs from three sites) [17]. No crabs were found in 2015–2016 along the south-eastern Awo River near one of the 2009 capture points (Nyalebbe HA). Overall, no active crab-associated *S. neavei* breeding was detected in 2015–2016 at the prospected locations.

Regarding non-crab *Simulium* larval breeding habitats, between 2015 and 2017, larval and pupal prospections documented breeding of *S. dentulosum* and *S. vorax* at several basins: 1) Awo (Kakoi River basin) in Logo HZ, Thedeja HA*: S. dentulosum*, *S. vorax*; and Nyarambe HZ, Nyalebbe HA: *S. vorax*; 2) downstream Kakoi River basin in Nyarambe HZ, Afoyo HA: *S. vorax*; 3) Kuda River basin in Angumu HZ, Kudieweka HA: *S. vorax*; Logo HZ, Draju HA: *S. dentulosum*, *S. vorax*; and Nyarambe HZ, Kpanyi HA: *S. vorax*; and 4) Koda River basin in Rethy HZ, Lokpa HA: *S. dentulosum*; and Rassia HA: *S. dentulosum* [17]. Breeding of several non-anthropophagic *Simulium* species was also recorded [17].

##### Adult female blackfly collections by informal “spot-check” HLCs

Informal HLCs undertaken in August–November 2017 and February–April 2018 identified two anthropophagic blackfly species within the Kakoi–Koda focus, namely, *Simulium (Anasolen) dentulosum* [63] and *S. vorax* Pomeroy [17, 49, 64]. Collections were made along the Kuda River basin across multiple HZs (HA): Angumu (Dabu), Logo (Draju), Nyarambe (Afoyo and Kpanyi) and Rethy (Rassia). Totals captured per HA were as follows: Afoyo 139 *S. dentulosum* and 21 *S. vorax*; Kpanyi 17 *S. dentulosum*; Dabu 2 *S. dentulosum* and 1 *S. vorax*; Draju 1 *S. vorax*; and Rassia 3 *S. dentulosum*.

A subset of the collected blackflies (155/161 *S. dentulosum* and 4/23 *S. vorax*) was processed individually for triplex real-time PCR (heads/bodies separated) to detect *O. volvulus* DNA. Among *S. dentulosum*, 30.3% were body-positive for *O. volvulus* (47/155; 95% CI: 23.3–38.3) and 11.0% were head-positive (infective; 17/155; 95% CI: 6.7–17.2). Four *S. dentulosum* flies were positive in both compartments, giving an overall prevalence of infection (in head and/or body) of 38.7% (60/155; 95% CI: 31.1–46.9). For *S. vorax*, 1/4 (25%) bodies and 0/4 (0%) heads were positive.

### 3.2 Geospatial analysis

An interactive ArcGIS web map aggregating epidemiological, entomological and land-cover layers (REMO/REA 2003–2015; skin-snip screening 2009–2011, 2015–2017, 2021–2023; anti-Ov16 surveys 2015–2021; HLC and breeding sites; forest loss 2001–2023; administrative boundaries; rivers/streams) is publicly available at https://arcg.is/1PDyO0. A README file is available in S1 Appendix Text A.

#### 3.2.1 Individual and spatial concordance of recent transmission indicators

Paired anti-Ov16 RDT and skin-snip microscopy results were available for children aged 3–10 years from the 2015 epilepsy case-control [45] and 2017 epilepsy cohort [47] studies (n = 83). Agreement between tests was high by simple proportion (94.0%), while chance-corrected agreement was moderate (κ = 0.52, 95% CI: 0.15–0.88; −1 to 1 scale), consistent with low prevalence. There was no evidence that the tests differed in overall positivity rates (mid-p McNemar p = 0.219). Using skin-snip microscopy as reference, anti-Ov16 RDT in the field had 42.9% sensitivity (95% CI: 9.9–81.6) and 98.7% specificity (95% CI: 92.9–100.0).

Anti-Ov16 seroprevalence in children aged 3–10 years sampled from the community (2016–2021; including unpublished data from the NSETHIO programme [30, 31]) and adolescent (12–17 years) microfilaria-positive skin-snip prevalence in 2021–2023 (second moxidectin trial screening) showed a coherent, geographically structured pattern (see interactive map https://arcg.is/nb45e1). Positive surveys were clustered along north-eastern Muda River tributaries of the Kuda River basin, and north-western tributaries of the Lebu River basin in Draju and Kanga HAs (Logo HZ), with 6/110 children seropositive for anti-Ov16 IgG4 and 40/1,010 adolescents testing as microfilaria-positive by skin-snip microscopy. Adjacent Kuda River tributaries upstream of the Muda River showed additional low-positive signals, with children’s serology yielding mixed results across nearby villages (4/127 seropositive), and adolescent microfilarial prevalence mirroring this pattern (27/908), producing a mosaic of zero to very low positivity in surveys conducted in Draju and along the borders with Ulyeko and Buu HA (Logo HZ).

In contrast, the north-eastern Kuda River branch (Kpanyi HA, Nyarambe HZ) showed no evidence of recent transmission, with 0/82 children seropositive and 0/344 adolescents skin-snip microfilaria-positive. Also, no evidence of recent transmission was detected in the Awo River basin or in the Kakoi River tributaries (including the sections of Suu and Ryeda rivers north of the Muda/Kuda), where only skin snips were undertaken (0/128; in Thedeja HA, Logo HZ).

Findings from the epilepsy case-control (2015) and the epilepsy cohort (2017) studies were consistent with this spatial pattern. Among 3–10-year-olds in the Kuda River basin (Draju), 10/43 skin snips and 2/14 anti-Ov16 tests were positive. Also, between the Koda and Loda River basins (Lokpa and Rassia HAs, Rethy HZ), 8/43 skin-snips and 3/18 anti-Ov16 tests were positive. Together, these data indicate that field anti-Ov16 RDT results in children reproduce the spatial gradient observed for adolescent skin-snips.

#### 3.2.2 Landscape change and tree canopy loss

Remote-sensing and field observations indicate substantial landscape transformation across the Kakoi–Koda focus [65]. Agriculture (e.g., coffee, rice, tobacco and cotton), logging and conflict-related population movements have contributed to progressive deforestation, already documented in western South Ituri by the late 2000s [65]. From 2001 to 2024, cumulative forest (and “only dense forest”) loss at the HZ level was 3.4% (39.5%) in Angumu, 4.4% (62.4%) in Logo, 4.0% (81.8%) in Mahagi, 4.7% (85.1%) in Nyarambe and 7.1% (12.8%) in Rethy. Dense forest loss was concentrated in 2000–2012 across Angumu, Logo, Mahagi and Nyarambe, with additional loss in 2017–2019 in Logo and in 2019 in Mahagi (S1 Appendix Fig A). In contrast, Rethy (with 96% of the dense forest biome in the focus) shows a more progressive pattern of dense forest clearance, accelerating since 2017.

At the HA level, among hyperendemic HAs identified through REMO (2003) or the 2009–2011 moxidectin trial skin-snip population screening, forest (and “only dense forest”) loss from 2001 to 2024 were: 1) Angumu HZ: 2.3% (12.2%) in Bessi, 4.1% (100%) in Dabu and 3.4% (40.4%) in Langa; 2) Logo HZ: 8.4% (89.6%) in Draju and 4.1% (73.8%) in Kanga; and 3) Nyarambe HZ: 2.5% (100%) in Afoyo and 6.5% (83.8%) in Kpanyi. In mesoendemic HAs, losses for forest (and “only dense forest”) were: 1) Angumu HZ: 1.3% (1.9%) in Aree and 4.5% (20.6%) in Musongwa; 2) Nyarambe HZ: 9.1% (100%) in Pundiga; and 3) Rethy HZ: 14.1% (34.4%) in Rassia. Data on dense forest cover and cumulative loss for other HAs within the 5 HZ of the Kakoi-Koda focus are available in the S1 Appendix Table A.

Canopy loss was spatially heterogeneous across the focus (Fig 4 and S1 Appendix Fig B). The highest losses were concentrated in the hyperendemic belt between Angumu, Logo and Nyarambe HZs, where dense forest has been extensively cleared. This includes the Awo River basin (74% deforested, mainly in Buu, Leo and Thedeja HAs: S1 Appendix Fig C), the Muda River tributaries in the Kuda River basin (78% deforested, largely in Draju HA and its borders with Kpanyi, Ambere and Kanga HAs: S1 Appendix Fig D), the northern Lebu River basin (92% deforested, around Kudieweka HA, adjacent to Kanga HAs: S1 Appendix Fig E), and the northern Koda and Loda River basins (83% deforested, in the Kpandroma-Wala-Lokpa HAs boundary: S1 Appendix Fig F). Small remnants of dense forest are located along the north-western Kakoi River tributaries (53% deforested, Langa HA: S1 Appendix Fig G). Larger mosaics of dense forest remain in the mid- and downstream Koda and Loda River basins (Rassia and Lokpa HAs, respectively), which have still undergone substantial loss (33% and 53% deforested, respectively: S1 Appendix Fig H), especially surrounding riparian (along the river bank) zones, potentially reducing suitable habitats for *S. neavei*. The Mount Aboro reserve in Uketha HA (bordering Rassia HA, Rethy HZ), also shows dense forest loss (13% deforested: S1 Appendix Fig H) over the period.

**Fig 4.**
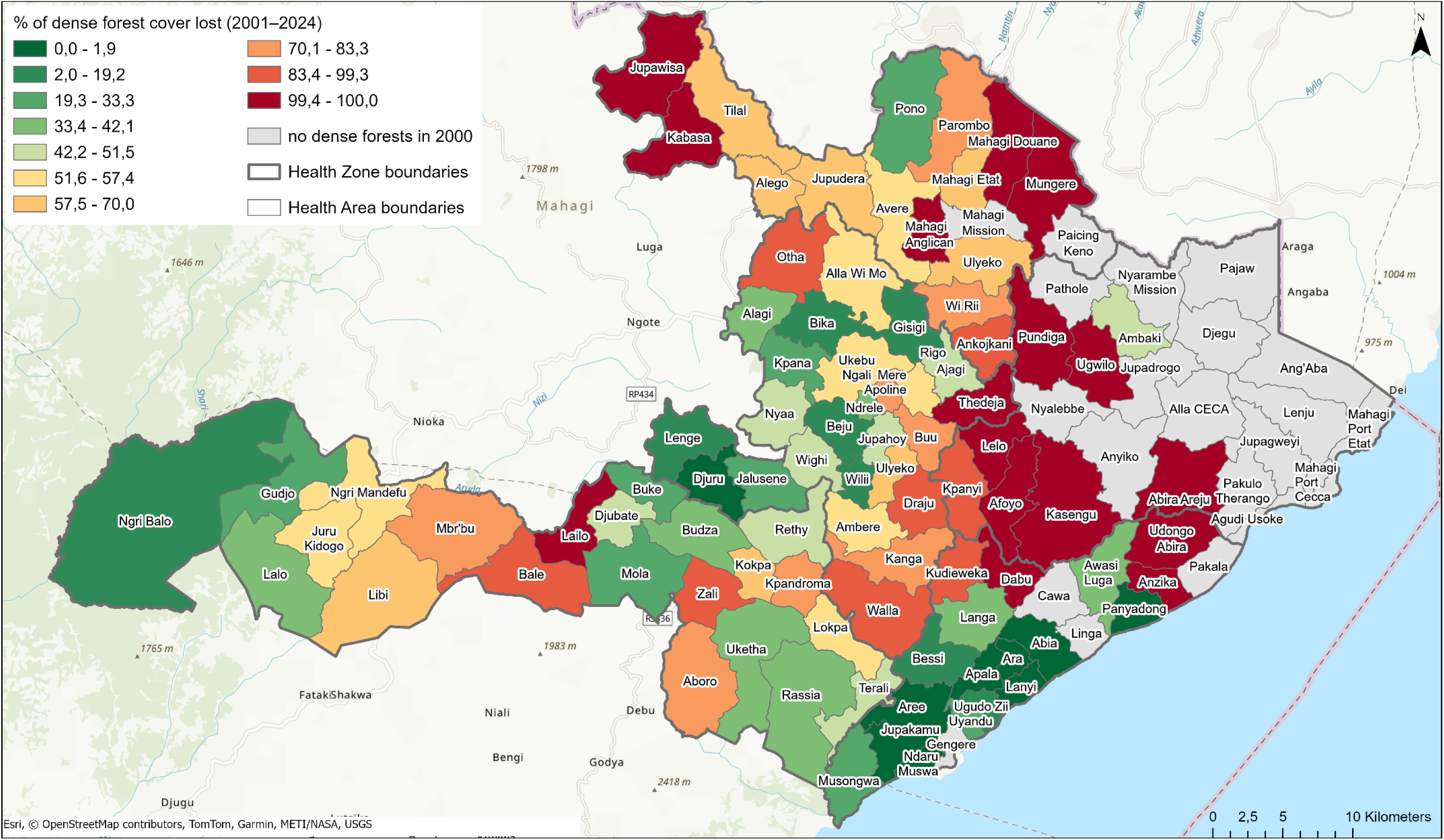
Percentage of dense forest cover lost between 2001 and 2024 in the Kakoi-Koda onchocerciasis focus. Health areas (black thin boundaries) are shaded by the percentage of dense forest (≥70% tree cover in 2000) that was lost by 2024, grouped into 10 quantile classes (from 0 to 100% loss). Health zone boundaries for Angumu, Logo, Mahagi, Nyarambe and Rethy are shown as bold grey lines. The map was produced in ArcGIS Pro (Esri, Redlands, CA, USA) using publicly available GRID3 COD – Health Areas v5.0 [27] and GRID3 COD – Health Zones v5.0 [28] boundaries and the Hansen Global Forest Change dataset (30-metre resolution; percent tree cover in 2000 and annual tree loss, 2001–2024) [33, 34].

## 4. Discussion

The Kakoi–Koda onchocerciasis focus in the DRC spans five HZs. Baseline nodule prevalence mapping (REMO, 2003) indicated hyperendemicity in parts of Logo, Nyarambe and Angumu HZs, with mesoendemicity in Mahagi and Rethy HZs. Across independent data streams spanning two decades, onchocerciasis transmission in the focus appears to have fallen sharply since the 2009–2011 (skin-snip) population screening for the Phase III moxidectin clinical trial [23, 24], which confirmed hyperendemicity in Logo and Nyarambe. Age (and sex) profiles of all-age anti-Ov16 surveys in 2015–2017 confirmed substantial cumulative exposure in parts of Logo and Rethy HZs, consistent with the baseline mapping.

By contrast, children-only (aged 3–10 years) anti-Ov16 seroprevalence from 2015 onward was uniformly low or zero in most surveyed HAs of Logo, Nyarambe and Rethy HZs that had been hyperendemic at baseline, with some positives mostly in older children (7–10 years), consistent with a reduction in the force of infection. Skin-snip screening corroborated this trajectory, with hyperendemic infection prevalence and intensity in 2009–2011 declining to hypoendemic levels by 2021–2023 in Draju and Kanga (Logo HZ), with the steepest relative declines in adolescents (the youngest group sampled using skin-snip microscopy). In Nyarambe (Kpanyi HA), where early sample sizes were small, prevalence in adults significantly fell from 72% to 3% in villages surveyed at both time points. In Rethy HZ, REMO categories changed from borderline hyperendemic (2003) to what would have been considered mesoendemic (2015), the latter result mirrored by microfilarial prevalence and intensity in studies among persons with epilepsy (2015). Spatially, residual transmission signals clustered along limited stretches of the Muda/Kuda and Lebu River basins (and possibly between Koda and Loda River basins). Other river basins, such as sections of Awo and Kakoi, showed no evidence of recent transmission.

The exact onset of the endemicity decline cannot be empirically dated due to the absence of earlier microfilarial data. However, in Draju (Logo HZ), the absence of anti-Ov16 seropositivity among young children aged 3–6 years in 2016 (0/70) indicates that transmission had already become very low by that time. By 2021–2023, infection prevalence and intensity had dropped to low levels in previously hyperendemic HAs in Logo and Nyarambe HZs, consistent with sustained reductions in the force of infection over more than a decade.

The age-specific community anti-Ov16 seroprevalence profile observed in Logo HZ in 2016, with very low seroprevalence in young children, followed by a steep rise between 7 and 30 years of age and a plateau thereafter (males and females analysed together), is qualitatively similar to model-predicted age patterns of microfilarial and anti-Ov16 prevalence after 10–15 annual CDTI rounds at around 70% coverage in a recent transmission-modelling study [66]. In those simulations, such profiles arise when transmission has been substantially suppressed for one to two decades due to CDTI. Although Logo HZ has not been under routine CDTI, analogous age patterns could result from a prolonged reduction in the force of infection driven by ecological change and vector species shifts and density declines. The marked decline in microfilarial prevalence and community infection intensity between the 2009–2011 and 2021–2023 moxidectin trial screenings is also broadly consistent with modelled trajectories for settings where transmission has declined over 15–20 years by 2021–2023 [66]. This congruence supports the hypothesis that *O. volvulus* transmission in Logo HZ has been decreasing since at least the early 2000s. Transmission-modelling analyses using the longitudinal microfilarial and anti-Ov16 seroprevalence data compiled here would be needed to test this hypothesis quantitatively.

Vector ecology provides a plausible mechanism for changing transmission patterns [17]. Baseline meso- and hyperendemic onchocerciasis clusters at the Kakoi-Koda focus lay predominantly along escarpment slopes between higher and lower elevations, consistent with rapids suitable for *Simulium* breeding (see Fig 2 and relief model of the focus in [17]). The *Simulium neavei* group is phoretically associated with freshwater crabs and breeds in shaded streams, where its immature stages attach to crabs for transport and feeding [67]. While *S. neavei* is sensitive to canopy opening and habitat disturbance [38, 68], *S. dentulosum* has been collected biting humans in the Ituri highlands and may act as a vector in more open habitats, with *S. vorax* possibly also contributing to transmission [17].

Historical information suggests that the *S. neavei* group occurred locally or in nearby Ituri rivers [69]. However, recent entomological work identified *Simulium dentulosum* and *S. vorax* as the anthropophagic species currently biting within the focus, with *O. volvulus* ND5-positive bodies (and for *S. dentulosum*, also heads), indicating parasite circulation along the Kuda River system in 2017–2018 [17]. In contrast, surveys since 2015 did not detect any active crab-associated *S. neavei* breeding in the prospected reaches (only one site with infested crabs was documented in Kakoi River in 2009) [17, 49], consistent with its apparent disappearance from the focus. This pattern, together with progressive deforestation and canopy opening, is compatible with reduced shaded-stream habitat and disruption of crab-associated breeding, similar to the disappearance of *S. neavei* observed after bush-clearing in a hypoendemic focus in Nyanza Province, Kenya [70]. In several Ugandan onchocerciasis foci, the disappearance of *S. neavei* has likewise been attributed to deforestation [71]. These observations are consistent with the hypothesis proposed by Post *et al.* [17] of a shift in the local vector community from *S. neavei* to more open-habitat species (*S. dentulosum* and *S. vorax*), with lower overall vectorial density and/or capacity. Additionally, since the early 2010s, sociopolitical instability and immigration of relatively wealthier households who are less engaged in farming (and other high-risk activities for *O. volvulus* transmission) may have further reduced human-vector contact [72].

CDTI histories were heterogeneous across the focus. Angumu and Rethy HZs had long-running CDTI programmes for onchocerciasis since, respectively, 2009 and 2012, and Nyarambe HZ implemented ivermectin and albendazole delivery from 2016 for lymphatic filariasis, all programmes reporting high therapeutic coverage. In contrast, no CDTI was delivered in Logo or Mahagi HZs. Beyond routine delivery, microfilaricidals had been provided to a small subset of residents recruited through moxidectin–ivermectin clinical trials (2009–2011; 2021–2026) [21, 24] and an ivermectin trial among persons with epilepsy (2017–2018) [55, 56]. Although the extent of these treatments was circumscribed (single dose in the first Phase III moxidectin trial; 1–3 annual doses or five biannual doses in the subsequent Phase IIIb moxidectin trials, and 1–3 doses in the ivermectin trial for epilepsy per person recruited in the trials’ arms) rather than population-wide, enrolment often focused on individuals with higher microfilarial loads (to assess drug effects on infection intensity or seizure outcomes). Therefore, these targeted treatments may have contributed to the area-wide decline observed, together with CDTI (in three of the five HZs) and decreasing human–vector contact linked to ecological change.

The marked onchocerciasis prevalence decline in Logo despite the absence of routine CDTI points towards a substantial role for ecological drivers, particularly canopy loss along streams, likely reducing suitable breeding habitat for *S. neavei* and constraining transmission to short river segments where anthropophagic *Simulium* species remain, but whose vector competence and ability to maintain transmission are uncertain. The concordance between very low-to-absent anti-Ov16 seroprevalence in children, decreasing adolescent and adult infection prevalence and intensity, and the contraction of transmission suggested by the evidence to the Muda/Kuda–Lebu rivers corridor (where CDTI has not been delivered) supports this interpretation. Future transmission modelling could account for both programmatic exposure (including the targeted trial doses) and ecological change when attributing the observed declines.

Two practical surveillance points emerge from our study. Firstly, anti-Ov16 RDTs performed in the field among children aged 3–10 years reproduced the spatial gradient seen in skin snips, despite diagnostic limitations (limited sensitivity and RDT inability to distinguish ongoing from past infection) [50]. In our paired analysis of skin-snip microscopy and anti-Ov16 RDT in children, the field RDT showed moderate sensitivity (43%, 95% CI: 10–82) and high specificity (99%, 95% CI: 93–100) against skin-snip microscopy. A similar diagnostic performance range has been reported for the same field-based RDT in Burkina Faso among 400 children aged 3–9 years (sensitivity = 60% and specificity = 94%, when compared to skin-snip microscopy) [73] and for all ages in an ivermectin-naïve setting in Gabon (sensitivity = 52% and specificity = 95%, when compared to skin-snip microscopy for 4,257 participants aged ≥5 years) [74]. These estimates do not reach the WHO diagnostic Target Product Profile (TPP), which calls for very high specificity (≥99.8%), and more moderate sensitivity thresholds (≥60% for elimination mapping and ≥89% for stopping CDTI decisions) [75].

Secondly, REMO/REA nodule palpation remains helpful to delineate baseline endemicity, particularly in areas of moderate and high transmission, but is less sensitive in areas of low transmission or to indicate transmission declines than microscopy or serology [18]. For example, in the Kakoi-Koda focus, nodule prevalence fell from hyperendemicity to mesoendemicity in Logo HZ between 2003 and 2015, and from high mesoendemicity to what would be considered hypo-/low mesoendemicity in Rethy HZ, but these shifts were less informative than anti-Ov16 seroprevalence and skin-snip prevalence trends.

### 4.1 Limitations

We integrated programme histories, REMO/REA, study-based and community serology and skin-snips, and entomology with geospatial layers, enabling triangulation across designs and time. This approach provides a comprehensive picture of the Kakoi–Koda focus but has several constraints. Firstly, several contributing studies were not designed to yield a population-representative sample for the focus as a whole; most had pragmatic or study-specific enrolment criteria (i.e., trial screening examining by skin-snip only those aged ≥12 years, and case–control and epilepsy cohort studies purposive sampling), limiting generalisability. We addressed this limitation by including previously unpublished community serology surveys among children in Logo and Nyarambe HZs conducted in 2018 and 2021, respectively, as indicators of recent/ongoing *O. volvulus* transmission. We further mitigated this limitation by synthesising information across independent sources and emphasising consistent patterns rather than single-study estimates.

Secondly, heterogeneity in study designs (trial screening, cross-sectional, case–control, cohort) and protocols (two versus four skin snips; microfilariae counted per snip or milligram of skin; differing age groups) limits precision and hampers direct comparisons of microfilarial intensity. The sensitivity of skin-snip microscopy increases with the number of snips and is reduced in light infections (e.g., in low-transmission or settings under CDTI), as suggested by modelling work [53]. Similarly, the Ov16 IgG4 RDT performed on whole blood has moderate and setting-dependent sensitivity [50, 76]. To minimise bias, we restricted formal comparisons within study design groups, reported 95% CIs and ranges, and used exact tests (Fisher/Clopper-Pearson) when data were sparse.

Thirdly, entomological findings are constrained by seasonal gaps, limited PCR testing of *S. vorax* for *O. volvulus* larvae, and incomplete spatial coverage (notably across parts of Kakoi, Awo, Kuda/Muda and Lebu rivers), which limit the understanding of the capacity of these simuliid species to sustain *O. volvulus* transmission. Nevertheless, *S. dentulosum* samples were confirmed to indicate infectivity (*O. volvulus* DNA in heads) within the focus [17], and *S. vorax* has demonstrated competence for *O. volvulus* in laboratory settings [64] and transmits *Onchocerca dukei* in cattle [17, 77], indicating that both species remain epidemiologically relevant.

Lastly, self-reported ivermectin intake in Kpanyi (very low among the moxidectin trial participants in 2021–2023) appears inconsistent with high CDTI coverage reports available in ESPEN for Nyarambe (80–84%) [20] and with the 2021 coverage among ivermectin-eligible children in Kpanyi (92%). While possible explanations include social desirability or reporting bias during screening and different sampling frames, this discrepancy warrants a targeted coverage verification survey.

### 4.2 Recommendations

Operationally, and in line with the WHO 2030 onchocerciasis elimination goal [1], risk-based surveillance should prioritise the Muda/Kuda and Lebu River basins and adjacent communities to determine whether CDTI should be initiated in Logo HZ and continued in the neighbouring HZs. Where ongoing transmission is verified, entomological monitoring should be considered to further characterise breeding sites, vector species, seasonal biting patterns, and infection/infectivity rates to guide interventions. Independent qualitative interviews with communities in Draju HA also identified the Kuda and Lebu lowland valleys as areas of highest current blackfly biting nuisance [31], independently highlighting these basins as priority areas for surveillance.

The assembled, standardised dataset accompanying this study may support transmission modelling to simulate prevalence changes in the focus [78]. In addition, blood-meal analysis could clarify host preferences of the current *Simulium* species found biting on humans and whether habitat change has altered human-vector contact, particularly in areas where flies may feed away from humans.

Although the case-control and cohort studies that contributed *O. volvulus* serology and skin-snip data were not designed to be representative of the population, as they focused on epilepsy, they were informative for tracking epidemiological trends and, together with the recent unpublished community child surveys and second moxidectin skin-snip trial, helped identify areas where transmission appears reduced but not fully suppressed. Leveraging such local studies and other health programmes, for example, integrating anti-Ov16 RDTs or opportunistic skin-snip surveys into ongoing research or service platforms, may offer a pragmatic, interdisciplinary approach to strengthen NTD surveillance and optimise the limited logistical and financial resources available to tackle them, especially as external funding declines [79, 80]. To maximise impact, datasets generated through such local studies and related health programmes should be shared openly to support planning and evaluation, as demonstrated in the depth of our analysis, which was possible only because previous data were accessible.

### 4.3 Conclusion

Epidemiological, entomological and geospatial evidence consistently indicate that *O. volvulus* transmission in the Kakoi–Koda focus has declined substantially since 2010. The most plausible explanation is the disappearance of *S. neavei*-compatible habitats and possible replacement by *S. dentulosum* and *S. vorax*. Programmatic effects (CDTI and small-scale trial treatments) likely reinforced these declines when and where delivered. Whether the current vector species can sustain endemically-stable transmission remains uncertain, as current evidence suggests that recent transmission is limited and spatially fragmented. Targeted, integrated surveillance focused on the identified sub-basins can be used to guide decisions on initiating/continuing or stopping CDTI and to support post-treatment surveillance. The standardised dataset assembled here provides a platform for onchocerciasis transmission modelling. In operational terms, anti-Ov16 RDTs performed in the field mirrored spatial patterns derived from skin-snip assessments, supporting their use as a complementary surveillance tool where the deployment of better diagnostics may not be feasible.

## Author contributions

**Luís-Jorge Amaral**: conceptualisation, data curation; formal analysis; funding acquisition; investigation; methodology; project administration; software; validation; visualisation; writing-original draft preparation; writing-review & editing. **Tony Ukety**: conceptualisation, data curation; funding acquisition; investigation; methodology; project administration; resources; writing-original draft preparation; writing-review & editing**. Jules Upenjirworth**: conceptualisation; funding acquisition; investigation; methodology; project administration; resources; writing-original draft preparation; writing-review & editing. **Deogratias U. Wonya’Rossi**: conceptualisation; writing-review & editing. **Michel Mandro**: conceptualisation; writing-review & editing. **Francoise Nyisi**: conceptualisation; writing-review & editing. **Pascal Adroba**: conceptualisation; writing-review & editing**. Wilma A. Stolk**: conceptualisation; funding acquisition; methodology; writing-review & editing**. Joseph N. Siewe Fodjo**: conceptualisation; writing-review & editing. **María-Gloria Basáñez**: conceptualisation; funding acquisition; methodology; resources; supervision; validation; writing-review & editing**. Anne Laudisoit**: conceptualisation; data curation; methodology; supervision; validation; writing-original draft preparation; writing-review & editing**. Robert Colebunders**: conceptualisation, data curation; formal analysis; funding acquisition; investigation; methodology; project administration; resources; supervision; validation; writing-original draft preparation; writing-review & editing.

## Data availability

The datasets generated during and/or analysed during the current study are freely available to other researchers. Please refer to the Supplementary S1 Appendix and S1 Dataset.

## Acknowledgements

The authors thank Dr Rory J. Post for his insightful advice and constructive feedback on earlier versions of the manuscript, and for his foundational work in delineating the Kakoi–Koda focus, which informed the conception of this study.

## Supporting information

**S1 Dataset**. Supplementary dataset containing the epidemiological data used in the analyses for the Kakoi–Koda onchocerciasis focus.

**S1 Appendix**. <u>Text A</u>: ArcGIS README file for the interactive map of epidemiological, entomological and landscape data for the Kakoi–Koda onchocerciasis focus. <u>Text B</u>: Anti-Ov16 ELISA and concurrent skin-snip microscopy shared by ESPEN (context surrounding the Kakoi–Koda focus). <u>Figures A–H</u>: Annual dense forest loss (2001–2024) by health zone and basin-level maps of dense forest in 2000 and dense forest loss (2001–2024) across the Kakoi–Koda focus (Awo basin; Muda tributaries of the Kuda basin; northern Lebu basin; northern Koda and Loda basins; north-western Kakoi basins; southern Koda and Loda basins and Mount Aboro). <u>Table A</u>: Dense forest cover in 2000 and cumulative loss (2001–2024) by health area in the five health zones of the Kakoi–Koda focus.

